# Major depression symptom severity associations with willingness to exert effort and patch foraging strategy

**DOI:** 10.1101/2024.02.18.24302985

**Authors:** Laura A. Bustamante, Deanna M. Barch, Johanne Solis, Temitope Oshinowo, Ivan Grahek, Anna B. Konova, Nathaniel D. Daw, Jonathan D. Cohen

**Author notes:** Corresponding author: Laura Ana Bustamante Washington University in St. Louis 1 Brookings Drive MSC 00334 St. Louis, MO, 63105.

## Abstract

**Background:** Individuals with major depressive disorder (MDD) can experience reduced motivation and cognitive function, leading to challenges with goal-directed behavior. When selecting goals, people maximize ‘expected value’ by selecting actions that maximize potential reward while minimizing associated costs, including effort ‘costs’ and the opportunity cost of time. In MDD, differential weighing of costs and benefits are theorized mechanisms underlying changes in goal-directed cognition and may contribute to symptom heterogeneity.

**Methods:** We used the Effort Foraging Task to quantify cognitive and physical effort costs, and patch leaving thresholds in low effort conditions (reflecting perceived opportunity cost of time) and investigated their shared versus distinct relationships to clinical features in participants with MDD (N=52, 43 in-episode) and comparisons (N=27).

**Results:** Contrary to our predictions, none of the decision-making measures differed with MDD diagnosis. However, each of the measures were related to symptom severity, over and above effects of ability (i.e., performance). Greater anxiety symptoms were selectively associated with *lower* cognitive effort cost (i.e. greater willingness to exert effort). Anhedonia and behavioral apathy were associated with increased physical effort costs. Finally, greater overall depression was related to decreased patch leaving thresholds.

**Conclusions:** Markers of effort-based decision-making may inform understanding of MDD heterogeneity. Increased willingness to exert cognitive effort may contribute to anxiety symptoms such as worry. Decreased leaving thresholds associations with symptom severity is consistent with reward rate-based accounts of reduced vigor in MDD. Future research should address subtypes of depression with or without anxiety, which may relate differentially to cognitive effort decisions.

## Background

### Goal-directed behavior in major depressive disorder (MDD)

Individuals with major depressive disorder (MDD) can experience challenges with goal-directed behavior, including reduced motivation due to symptoms such as apathy, anergia, and anhedonia, as well as reduced cognitive function. Decisions about which goals to pursue and which actions to take to achieve them, can be understood in terms of costs and benefits. People maximize ‘expected value’ by selecting actions that maximize potential reward while minimizing associated costs. Effort-based decision making involves minimizing cognitive and physical effort costs (Rigoux and Guigon, 2012; Salamone, Correa, Yang, Rotolo, and Presby, 2018; Shenhav, Botvinick, and Cohen, 2013; Shenhav et al., 2017; Walton, Rudebeck, Bannerman, and Rushworth, 2007), as well as the opportunity cost of time (often also emphasized in value-based decision-making, Constantino and Daw, 2015).

Under effort- and value-based decision-making disruption accounts of MDD symptoms, changes in goal-directed behavior in depression can come from multiple causes, for example, differences in representing either the benefits or the costs of potential actions. The present study focuses on cognitive and physical effort costs, as well as opportunity costs (i.e., reward rate), to understand how differences in these components of goal-directed behavior relate to clinical features of MDD.

Both cognitive and physical effort-based decision-making have been reported to differ in MDD, though findings have been mixed. MDD has been associated with decreased willingness to exert cognitive effort relative to comparison groups in some studies (Ang, Gelda, and Pizzagalli, 2023; Vinckier et al., 2022; Westbrook et al., 2022) though not in others (Barch et al., 2023; Tran, Hagen, Hollenstein, and Bowie, 2021). Willingness to exert physical effort has been found to be decreased in MDD relative to comparison groups in some studies (Isabel M. Berwian et al., 2020; Cléry-Melin et al., 2011; Treadway, Bossaller, Shelton, and Zald, 2012; Vinckier et al., 2022; Wang et al., 2022; X.-H. Yang et al., 2014; Zou et al., 2020), though not in others (Cathomas et al., 2021; Sherdell, Waugh, and Gotlib, 2012; Tran et al., 2021; Wang et al., 2022; X. Yang et al., 2021).

### Dissociating between cognitive and physical effort costs

Both cognitive and physical effort-based decision-making appear to be associated with MDD features and may underlie certain MDD symptoms, though findings have been mixed. Importantly, MDD is highly heterogeneous in terms of variation in symptom domains and severity across individuals. This may contribute to inconsistent findings with respect to diagnostic group differences and associations with clinical features. MDD presentation encompasses many different symptoms, and decision-making mechanisms have many different components (including reward sensitivity, effort costs, task ability). Gaining traction on mechanistically informed treatments will require precise computational measures of decision-making to tease apart their specific relationships to precise symptom measures.

Initial studies measuring both effort types within-participants found differential relationships between cognitive and physical effort decisions and symptoms (Tran et al., 2021; Vinckier et al., 2022), suggesting potential applications to characterizing MDD heterogeneity. Studies measuring each effort type separately have reported decreased willingness to exert physical effort associated with symptom severity (i.e., anhedonia) (Sherdell et al., 2012; Tran et al., 2021; X.-H. Yang et al., 2014), while others reported no relationship to symptom severity (e.g., not related to depression, anhedonia, apathy, (Ang et al., 2023; Barch et al., 2023; Hershenberg et al., 2016; Vinckier et al., 2022). For cognitive effort, some studies report associations with symptom severity (i.e., global functioning, Tran et al., 2021; Westbrook et al., 2022) while others do not (e.g., not related to depression, anhedonia, apathy, Ang et al., 2023; Barch et al., 2023; Hershenberg et al., 2016; Vinckier et al., 2022). It remains unclear which symptoms map onto which component decision processes, and how shared or distinct these mappings are between cognitive and physical effort.

### Hypothesized symptom relationships

Multiple symptom MDD domains have been proposed to relate to value- and effort-based decision-making. Subjective reward rate, reflecting the opportunity cost of time, is proposed to drive the vigor of actions, represented via midbrain dopamine tone (Niv, Daw, Joel, and Dayan, 2007). Drawing on this, Huys, Daw, and Dayan (2015) proposed that physical anergia and psychomotor slowing symptoms of depression may be caused by reduced subjective reward rate representations. Relatedly, reduced willingness to exert physical effort to obtain rewards has been proposed as a mechanism underlying anhedonia and apathy symptoms (Cooper, Arulpragasam, and Treadway, 2018; Husain and Roiser, 2018; Pessiglione, Vinckier, Bouret, Daunizeau, and Le Bouc, 2018). Grahek and colleagues (2019) proposed that changes in motivational processes, resulting in reduced willingness to exert cognitive effort, may underlie, in part, reduced cognitive function associated with MDD, challenging the standard assumption that reduced cognitive function reflects reduced cognitive control capacity (Millan et al., 2012; Rock, Roiser, Riedel, and Blackwell, 2014; Snyder, 2013). This is of particular importance because reduced cognitive function in MDD contributes to disability (Jaeger, Berns, Uzelac, and Davis-Conway, 2006) and often does not improve with otherwise effective anti-depressant treatments (Halahakoon and Roiser, 2016; Rosenblat, Kakar, and McIntyre, 2016). By the cognitive effort-based decision-making account, interventions to improve cognitive function would focus on boosting motivation and target willingness to engage control, rather than cognitive control ability (e.g., computerized cognitive training) as suggested by the reduced capacity account.

The variable prevalence of these symptoms across studies may contribute to mixed findings. For example, reduced motivation may be minimal or absent in some individuals with MDD (Ang, Lockwood, Apps, Muhammed, and Husain, 2017; Nakonezny, Carmody, Morris, Kurian, and Trivedi, 2010). In addition, certain symptom domains of depression may show a differential relationship to effort relative to others. Anxiety (the most common MDD comorbidity, Kessler et al., 1996) symptoms such as rumination and worry may require cognitive effort (e.g., sampling for replay and planning (Bedder, Pisupati, and Niv, 2023) and anxiety has been related to increased effortful model-based planning (Gillan, Kosinski, Whelan, Phelps, and Daw, 2016). Anxiety has also been linked to increased cognitive effort exertion to maintain performance in the face of increased attentional demands posed by threat-related stimuli (Eysenck, Derakshan, Santos, and Calvo, 2007). Additionally, social anxiety may be associated with enhanced motivation to exert cognitive effort in social contexts (such as a psychology experiment, Hunter, Bornstein, and Hartley, 2018). It therefore may be important to account for anxiety heterogeneity and relate cognitive effort-based decision making to specific depression symptom expression profiles (or subtypes), as some have suggested (Gagne, Zika, Dayan, and Bishop, 2020; Lynch, Gunning, and Liston, 2020).

### Experiment overview

The goal of the current study was to quantify multiple components of effort-based decision-making and decompose their contributions to MDD symptom expression profiles. Each component measured (cognitive and physical effort cost, cognitive task ability, subjective opportunity cost of time) has been proposed as an underlying mechanism of specific MDD symptoms. To test these accounts, we had participants who met diagnostic criteria for MDD (most in-episode) and demographically matched comparison participants with no psychiatric diagnoses complete the cognitive and physical Effort Foraging Task (Bustamante et al., 2023). To our knowledge, all previous MDD studies used explicit tasks in which participants choose between low-effort/low-reward and high-effort/high-reward options. We hypothesized that the Effort Foraging Task, which measures effort avoidance more implicitly by inferring the cost of effort from foraging behavior, would be less contaminated by demand characteristics that bring about changes in what participants value about effort (e.g., try to please the experimenter, Orne, 1962). Reduced control over demand characteristics in explicit tasks, in turn, may contribute to mixed findings. While we hypothesized the Effort Foraging Task would yield more valid findings regarding the relationship between MDD and willingness to exert effort, it could also reasonably be the case that this task may tap into distinct aspects of effort-based decision making than the tasks used in the extant literature (e.g., implicit versus explicit processing).

In the Effort Foraging Task, participants choose between harvesting a depleting patch, or traveling to a new patch, which is costly in time and effort. Participants completed the 3-Back level of an N-Back working memory task (Nystrom et al., 2000) in the high cognitive effort condition, the 1-Back level in low cognitive effort condition. Participants completed a larger number of rapid keypresses in the physical high effort condition and a smaller number of presses in the low physical effort condition. Analyses focused on ‘exit thresholds’, the reward value at which the participant decided to exit the current patch. The exit threshold reveals the point of equivalence in the tradeoff between the cost of harvesting with diminishing rewards and the cost of traveling to a new patch, and this is captured by a foraging-theory model (the Marginal Value Theorem, MVT, Charnov, 1976). According to the MVT, exit thresholds should reflect subjective average reward rate (i.e., opportunity cost of time), and this comports with human behavior (Constantino et al., 2017; Constantino and Daw, 2015; Lenow, Constantino, Daw, and Phelps, 2017). Based on this patch-leaving behavior, a foraging-theory-based computational model quantified individual differences in the ‘cost’ of effort. The longer a participant delayed leaving the patch in high versus low effort conditions (i.e., relatively lower the exit threshold) the larger their inferred effort cost parameter. Average exit thresholds in low effort blocks were used to assess overall foraging strategy, putatively reflecting subjective opportunity cost of time. Lastly, we assessed effortful travel task ability using accuracy and reaction times. We aimed to tease apart the influences of each of these effort decision-making components on clinical features of MDD by examining (i) diagnostic group differences, (ii) associations with overall depression severity, and (iii) associations with symptom severity.

Based on previous findings and theoretical work we predicted cognitive effort costs would be increased in the MDD group and related to cognitive function symptoms (i.e., subjective cognitive complaints relative to baseline, Grahek, Everaert, Krebs, and Koster, 2018; Grahek, Shenhav, Musslick, Krebs, and Koster, 2019). Based on previous findings with explicit tasks, we predicted physical effort cost would be increased in the MDD group and related to anhedonia (Sherdell et al., 2012; Tran et al., 2021; X.-H. Yang et al., 2014). Following Huys et al. (2015) we hypothesized that the MDD group would differ in foraging strategy, exhibiting lower exit overall thresholds, and that this would relate to physical anergia/slowing.

## Methods

### Study overview

#### Participants

97 participants volunteered for the study and gave informed consent as approved by the Rutgers University Institutional Review Board (67 MDD, mean=26.9 years, SD=11.1, 18-61; 30 comparison, mean=27.1 years, SD=9.64, 19-59, further details in S.I. section 1, Figure S.I. 1). Groups were matched on key demographic variables (S.I. section 1.2). We oversampled MDD participants to maximize power to detect continuous symptom relationships to task behavior within this group. The comparison sample size was then selected to be adequate to detect group differences in behavior. Power analysis indicated that we could detect a medium effect size for group differences and symptom relationships with 80% power (S.I. section 2). All participants completed a detailed clinical assessment in session 1, but 7 MDD and 3 comparison participants opted not to return. Clinical symptom ratings and self-report analysis included data from the 60 MDD and 27 comparison participants who returned for the second (task) session. 50 MDD participants were currently depressed (43 with task data), 6 were in partial remission and 4 were in full remission (all with task data, details in S.I. section 3). 32 MDD participants used psychotropic medication while 28 did not.

#### Clinician ratings and self-reports

The Structured Clinical Interview for DSM-5 (First, 2015) confirmed assignment of MDD and/or co-morbid anxiety diagnosis in the MDD group (and absence of exclusionary diagnoses in both groups, S.I. section 1), as well as whether participants were currently depressed (or in partial or full remission). Depression symptoms were assessed using the Hamilton Depression Rating Scale (HAMD, Hamilton, 1960), Brief Psychiatric Rating Scale (Overall and Gorham, 1962), MGH Cognitive and Physical Functioning Questionnaire (Fava, Iosifescu, Pedrelli, and Baer, 2009), Patient Heath Questionnaire-9 (PHQ-9, Kroenke, Spitzer, and Williams, 2001), Generalized Anxiety Disorder-7 (Spitzer, Kroenke, Williams, and Löwe, 2006), Snaith–Hamilton Pleasure Scale (Nakonezny et al., 2010), Apathy Motivation Index (Ang et al., 2017), Adult Temperament Questionnaire Effortful Control subscale (Evans and Rothbart, 2007), and Need for Cognition scale (Cacioppo, Petty, and Kao, 1984, S.I. section 4, Table S.I. 1).

Symptom severity was measured using confirmatory factor analysis to combine clinician-rated and self-report measures of the following symptoms: anhedonia, anxiety, appetite symptoms, behavioral apathy, emotional apathy, social apathy, cognitive function symptoms, depressed mood/suicidality, and physical anergia/slowing, as well as for trait effortful control and need for cognition (see S.I. section 5, Table S.I. 2, and Table S.I. 3). Items were assigned based on what each scale was validated to measure. Assigned items were z-scored and averaged to compute a symptom severity score in the MDD group only. Items with an inter-item correlation below 0.2 were eliminated (multilevel package, item.total function, Bliese, Chen, Downes, Schepker, and Lang, 2022). Internal consistency was computed using Cronbach’s alpha (Table S.I. 2, ltm package, cronbach.alpha function, Rizopoulos, 2006). Factors with alpha*<*0.6 were excluded from further analysis (i.e., emotional apathy, appetite symptoms). The resulting items were then applied to compute confirmatory factor scores for (1) MDD only, which was the focus of our analysis, and (2) all participants to test generalizability of effects.

### Effort Foraging Task

In the Effort Foraging Task participants harvested apples in virtual orchards (Figure S.I. 2, as described in Experiment 2 of Bustamante et al. (2023)). On each foraging trial the participant visits a ‘patch’ which can be harvested by pressing the down arrow key once to yield rewards (apples, converted to a monetary bonus to be incentive compatible). The marginal return decreases with each successive harvest. The initial reward from a patch was drawn from a normal distribution *N* (15, 1) and subsequent harvests were a product of the previous reward and a decay rate (drawn from a distribution beta distribution, *β*(14.90873, 2.033008), mean=0.88). The smallest reward for harvesting was 0.5 apples and participants were not prevented from persisting in extracting 0.5 apples from the patch. At any point the participant can travel to a new patch by pressing the right arrow key, which has replenished rewards, but it takes time and effort to travel there (Figure S.I. 3). The cognitive effort manipulation was the N-Back working memory task (1- and 3-Back levels, also used in Tran et al., 2021; Westbrook et al., 2022). The physical effort manipulation was rapid key pressing (also used in Isabel M. Berwian et al., 2020; Tran et al., 2021; Treadway et al., 2012; Treadway, Buckholtz, Schwartzman, Lambert, and Zald, 2009; Wang et al., 2022; X.-H. Yang et al., 2014; X. Yang et al., 2021) with the non-dominant pinky finger (50% or 100% of an individually calibrated maximum). Patches were presented block-wise in counterbalanced order (S.I. section 6). Blocks varied only in their (explicitly instructed) effort requirement (timing held constant, environment specifications in Table 1). Reaching a new patch was not dependent on performance. Participants had to reach a performance criterion during training to begin foraging (training and instructions in S.I. section 7).

**Table 1:** Foraging environment parameters and results of best threshold simulation. A: column 1: environment parameter, column 2: value. Participants completed eight 7-minute blocks. B: Best exit threshold policy identified in Bustamante et al. (2023), rows indicate best threshold from simulation, reward rate achieved with best threshold (apples per second), mean and standard deviation of the number of harvests it took to reach the best threshold.

| **A. Environment parameter** | **Value** |
| --- | --- |
| *Harvest time* | 2 seconds |
| *Travel time* | 20 seconds |
| *Cognitive Travel Tasks* | 1-Back, 3-Back (10 trials) |
| *Physical Travel Tasks* | Larger (100% max), Smaller (50%) number of presses |
| *Block duration* | 7 minutes |
| *Number blocks per condition* | 2 |
| *Number of conditions* | 4 |
| *Total Number of blocks* | 8 |
| *Initial reward* | $\mathcal{N}(15, 1)$ |
| *Decay rate ($\kappa$)* | $\beta(14.13, 2.03)$, mean = 0.88 |
| **B. Best threshold simulation** | **Value** |
| *Best threshold ($\rho$, apples)* | 4.65 |
| *Reward rate (apples/second)* | 2.32 |
| *Number of harvests $\mu$* | 9.95 |
| *Number of harvests $SD$* | 2.01 |

We followed a subset of exclusions validated in Bustamante et al. (2023) that most impede estimates of effort costs (S.I. section 8). Participants were excluded if they missed the response deadline on many foraging trials (1 MDD participant excluded missed 49.5%) or had very few exit trials (1 MDD participant was excluded for 1 high effort physical and 3 high effort cognitive exits, 1 MDD participant was excluded from cognitive effort analyses for 2 cognitive high effort exits). The final sample included in behavioral analyses was 52 MDD participants (53 MDD participants for physical effort) and 27 comparison participants. We confirmed there were no diagnostic group differences among participants included in task-based analyses.

### Marginal Value Theorem (MVT) model

The MVT predicts a forager should leave a patch when the instantaneous reward rate falls below the long-run average (Charnov, 1976; Constantino and Daw, 2015). Travel costs were estimated using a hierarchical Bayesian logistic model. For each trial (*t*), the model compared the expected reward on the next harvest (*R_e,t_*, Eq. (1)) against the condition-specific exit threshold (*ρ*_condition_. Expected reward was based on the last reward multiplied by the mean depletion rate (*kappa*). The first harvest of a patch, being forced, was excluded from analysis.

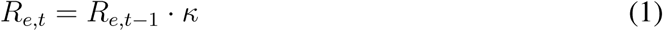

Using the difference of these to determine whether to harvest (1) or exit (0) via a softmax function (with inverse temperature, *β*, Eq. (2)).

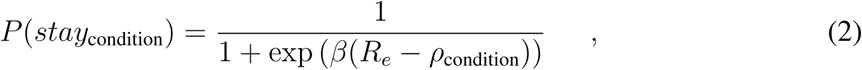

The cost of travel in high effort blocks (*c*_high effort_) was expressed as the marginal increase in cost of travel (*c*_low effort_ + *c*_high effort_) from low effort blocks, to control for any biases common to both conditions (e.g., variation in overall exit thresholds). Exit thresholds (*ρ*) were taken as fixed per-condition, determined by the total rewards (∑ *r*), total amount of time (number of harvest periods, *T* =condition duration/harvest time) and total travel costs (∑*c*, sum over total times travelled in a condition) across all blocks of a condition.

where,

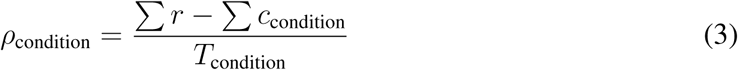

Individual and group-level parameters were estimated using Markov Chain Monte Carlo sampling (using cmdstanr, Stan, 2021). We used trace plots, R^^^ diagnostic statistics, and posterior predictive checks to assess model fit (S.I. section 9). We compared goodness of fit between diagnostic groups with an unpaired t-test on participants’ log posterior likelihoods (S.I. section 10). We compared participants’ overall exit thresholds to the best threshold found by simulation (S.I. section 12).

For model-agnostic analyses, we used linear mixed-effects regression to predict log transformed expected reward (Eq. (4)) on exit trials by an intercept term and effort level term separately for each effort type.

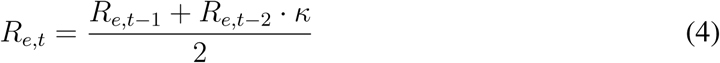

The first harvest of a patch was excluded from analysis, and on the second harvest of a patch we used the last reward multiplied by the depletion rate.

### Analysis overview

All subsequent analyses used point estimates (mean) from participant-level effort costs and applied them in frequentist tests to control for potential confounding variables and conduct multiple-comparison corrections. All analyses were conducted in the R language (many using the stats package, RCoreTeam, 2015). The HAMD total score was used to assess major depressive episode severity in the past week (herein ’overall depression’). We verified that results matched using self-reported depression (PHQ-9) due to concerns with HAMD validity (Bagby, Ryder, Schuller, and Marshall, 2004; Gibbons, Clark, and Kupfer, 1993; Ma et al., 2021). Our focus was on MDD symptom severity (z-scored for all analyses). To ensure remitted status was not driving key effects, we repeated all analyses zooming in on the current depressed group (excluding remitted participants). To test if effects were generalizable across the sample, we repeated analyses zooming out to all participants. We verified that no key results differed when controlling for psychotropic medication use (binary variable).

#### Diagnostic group differences

We tested for diagnostic group differences in cognitive or physical effort costs, controlling for high-effort task performance (3-Back D’ or % larger number of presses completed), years of education, age, and BMI (body mass index, for physical effort) using linear regression. To confirm hierarchical shrinkage did not bias results, we also fit group differences for all parameters directly within the MVT model (S.I. section 9). Additionally, we tested for a fatigue-like effect emerged within a block, and whether this differed by group (S.I. section 11).

#### Symptom associations with model parameters

Within the MDD group, we tested overall depression severity effects on cognitive or physical effort costs using linear regression, controlling for years of education, age, BMI (for physical effort) and high-effort task performance. Therefore, observed symptom associations are over and above effects of travel task ability. Next, we decomposed overall depression effects on effort costs into specific symptoms in a series of regression models. Because of mutual correlations between symptoms, we used multiple comparisons correction within a series of symptom models (FDR, 7 tests for each effort cost). We conducted a comparison of correlations to confirm specificity of observed relationships to effort type (cocor package) (Diedenhofen and Musch, 2015; Meng, Rosenthal, and Rubin, 1992). Because we allowed trial-wise variation to be captured by the inverse temperature parameter, we tested whether this parameter was correlated with symptom severity within the MDD group controlling for age and years of education.

#### Additional task measures

Participants may have differed in their ability to complete the required effort, which could confound decision-making differences and/or relate to symptoms. This is especially relevant for the cognitive task, which was not calibrated to individual ability. We addressed this by controlling for performance in symptom regression. We also examined whether performance was associated with (i) diagnostic group, (ii) overall depression, and (iii) symptom domains (S.I. section 13).

Some depression symptoms are theorized to arise from reduced subjective reward rate representations (Huys, Daw, and Dayan, 2015), which decrease vigor (Niv et al., 2007). We tested whether overall exit thresholds (from low effort conditions, which were least confounded by effort sensitivity) were associated with (i) diagnostic group, (ii) overall depression, or (iii) symptom domains (S.I. section 13.2).

## Results

### Demographic and clinical characteristics

Diagnostic groups were matched on gender, race, age, parental education, household income, and childhood income (also within participants included in behavioral analyses, Table S.I. 4). The comparison group had more years of education than the MDD group (Table S.I. 4), so it was included as a covariate in all analyses. Depression severity varied widely in the MDD group (Figure S.I. 4). The MDD group scored higher on all symptom domains except emotional apathy. Need for cognition did not differ between groups, while effortful control was higher in comparisons (Figure S.I. 5).

### Sensitivity to effort manipulations

On average, participants avoided effort (group-level posterior effort costs non-overlapping with zero, Table S.I. 5). The model converged (R^^^ *<*1.035 for all parameters, Figure S.I. 6) and the observed probability of harvesting, across participants and trials, fell within the posterior predictive distribution (*pd>*0.384). Simulated data recapitulated the empirical group-level change in exit threshold and overall exit threshold (Figure S.I. 7) as well as the probability of exiting by expected reward level relative to individual overall exit thresholds (Figure S.I. 8). We found no conclusive evidence for or against a correlation between cognitive and physical effort costs (mean=0.053, 95% HDI=-0.240, 0.345, Table S.I. 5, discussed in the context of Experiment 2, Bustamante et al. (2023), in S.I. section 14).

### Effort costs relationships to clinical features

#### Diagnostic group differences

We predicted effort costs by diagnostic group, controlling for high-effort task performance, education, age, and BMI (for physical effort). There were no group differences in either effort cost (Figure 1, cognitive: *p>*0.70, physical: *p>*0.77), even when controlling for psychotropic medication use (Table S.I. 6), and excluding remitted MDD participants (cognitive: *p>*0.47, physical: *p>*0.97). To ensure shrinkage in the hierarchical model did not obscure a group difference, we directly fitted diagnostic group differences for all MVT model parameters and found no differences in any of the parameters, even when excluding remitted MDD participants (Table S.I. 7). There were no group differences in the model-agnostic measure of effort sensitivity (S.I. section 16, Figure S.I. 9), nor in fatigue-like effects (S.I. section 17). We computed the log posterior likelihoods per participant and found no significant difference between diagnostic groups, suggesting comparable goodness of fit (Figure S.I. 7, *t*=0.56, *df* =36.28, *p>*0.58). Consistent with prior samples we found a minority of participants who had negative effort costs, consistent with effort seeking, but no significant difference between the groups (S.I. section 15).

**Figure 1:**
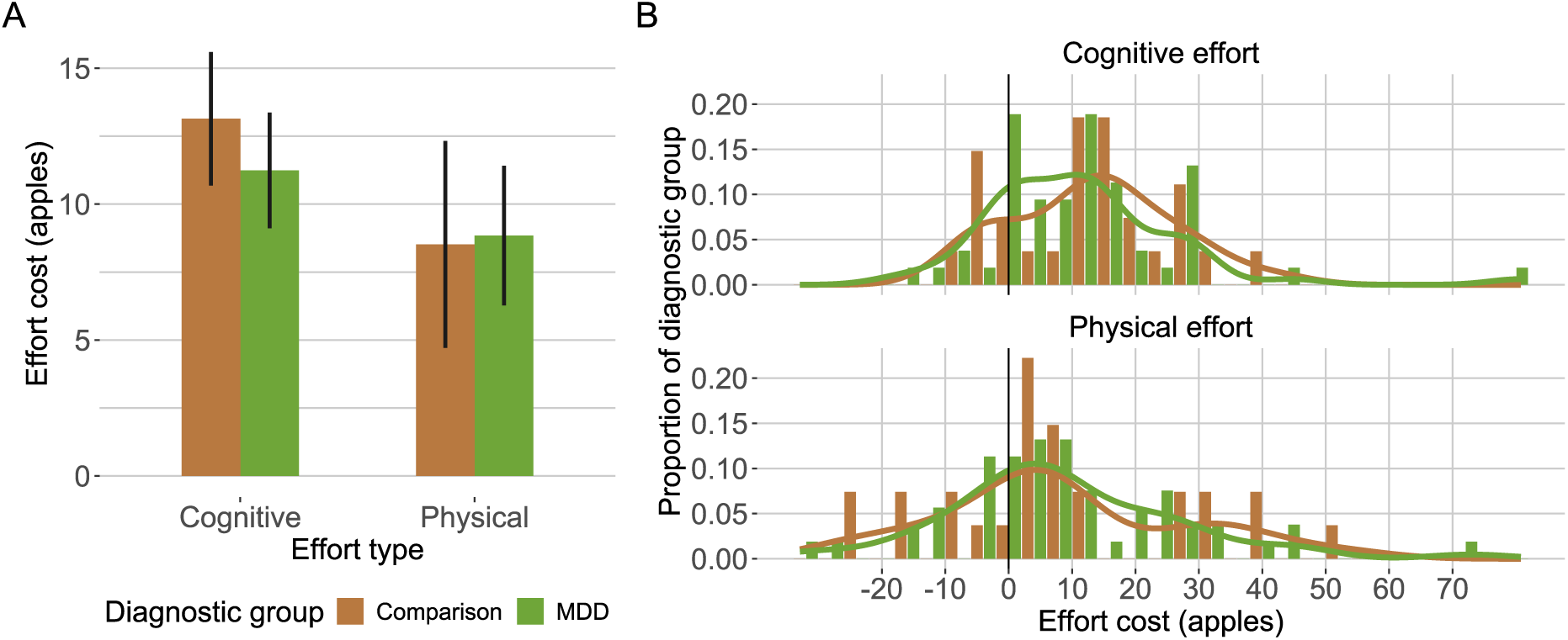
Effort cost by diagnostic group and effort type. Left panel: mean and standard error of the mean of individual differences in effort cost (y-axis) by effort type (x-axis). Right panel: individual differences histograms, x-axis indicates effort cost (larger values indicate more effort avoidance), y-axis indicates proportion of diagnostic group.

#### Overall depression severity

Surprisingly, overall depression severity was associated with *decreased* cognitive effort cost (*p<*0.030, Figure 2, Table 2). We found no reliable association with physical effort costs (Table 2) however, the correlation magnitudes between the two types of effort were not significantly different (*z*=-1.42, *p*=0.156). Results were maintained when using self-reported depression (PHQ-9) as the overall severity measure (Table S.I. 8), and when controlling for medication use (Table S.I. 6). However, the cognitive effort cost relationship was not maintained when restricting the analyses to current MDD participants (Table S.I. 9). Additionally, we found no reliable association with inverse temperature (Table S.I. 10). Next, we identified which symptom domains contributed to the cognitive effort cost relationship to overall depression, and whether physical effort cost was related to any symptom domain.

**Figure 2:**
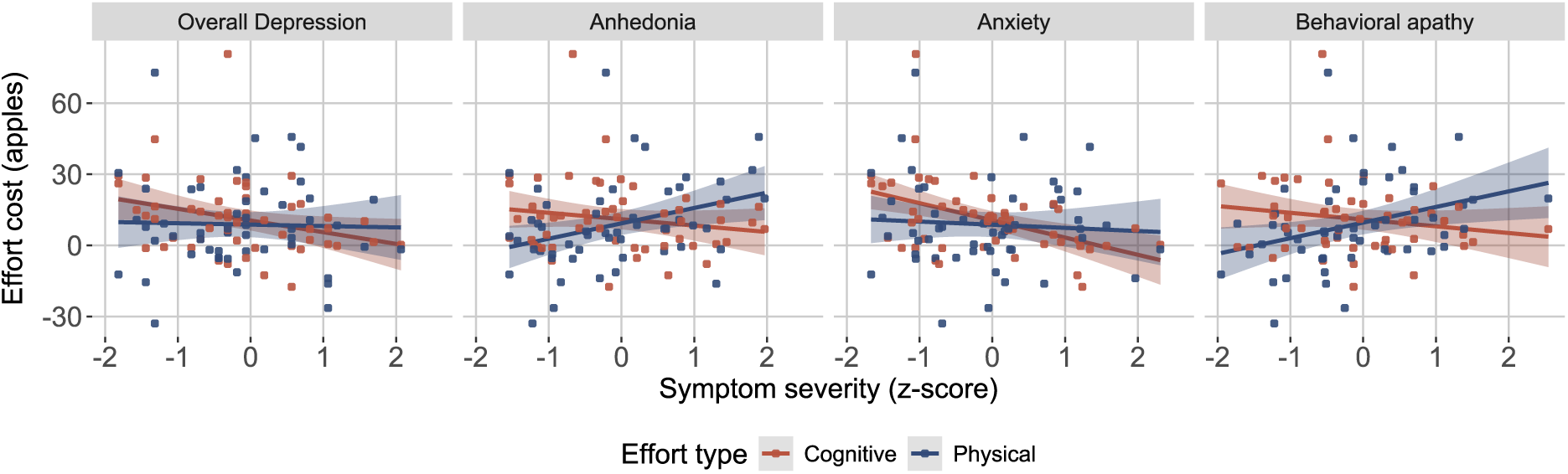
Relationship of individual MDD symptom domains with overall exit threshold (MDD group only). A: No diagnostic group differences (x-axis) in overall threshold (y-axis, apples, estimated from low effort conditions). Bar indicates group means, error bars indicate standard error of the mean, points indicate mean overall exit threshold per participant (i.e., random effects coefficients from linear regression model). B: Lower overall exit threshold (y-axes) were significantly related to overall depression severity (x-axes, Hamilton Depression Rating Scale Total z-score). Dashed line indicates best threshold policy, linear regression line for MDD group only.

**Table 2:** Symptom effort cost regressions (MDD group only). (A) Predicting cognitive effort cost from overall depression severity, and each symptom domain, controlling for cognitive task performance (3-Back D’) years of education and age. (B) Predicting physical effort cost from overall depression severity, and each symptom domain, controlling for physical task performance (% larger number of presses completed), BMI, years of education and age. (C) Predicting cognitive effort cost from each symptom domain, controlling for anxiety, cognitive task performance (3-Back D’) and age (* indicates *p<*0.05, FDR correction within symptom models). *R*^2^ and adjusted *R*^2^ displayed for significant models. All variables were scaled as input to the regressions.

| Symptom | Estimate | SE | *t* | *p* | *p*<sub>adjusted</sub> | *R*<sup>2</sup> | adj. *R*<sup>2</sup> |
| --- | --- | --- | --- | --- | --- | --- | --- |
| **A. Cognitive effort cost, MDD** |  |  |  |  |  |  |  |
| Overall depression | -0.37 | 0.16 | -2.24 | 0.030* |  | 0.120 | 0.045 |
| Anhedonia | -0.17 | 0.15 | -1.18 | 0.245 | 0.286 |  |  |
| Anxiety | -0.50 | 0.13 | -3.70 | 0.001 | 0.004* | 0.246 | 0.182 |
| Behavioral apathy | -0.19 | 0.15 | -1.28 | 0.205 | 0.286 |  |  |
| Social apathy | 0.01 | 0.15 | 0.06 | 0.950 | 0.950 |  |  |
| Cognitive function symptoms | -0.30 | 0.14 | -2.08 | 0.043 | 0.150 |  |  |
| Depressed mood/suicidality | -0.26 | 0.17 | -1.56 | 0.126 | 0.253 |  |  |
| Physical anergia/slowness | -0.22 | 0.15 | -1.48 | 0.145 | 0.253 |  |  |
| **B. Physical effort cost, MDD** |  |  |  |  |  |  |  |
| Overall depression | 0.04 | 0.17 | 0.21 | 0.833 |  |  |  |
| Anhedonia | 0.42 | 0.14 | 2.93 | 0.005 | 0.035* | 0.193 | 0.103 |
| Anxiety | -0.04 | 0.15 | -0.26 | 0.793 | 0.793 |  |  |
| Behavioral apathy | 0.39 | 0.15 | 2.71 | 0.010 | 0.035 | 0.173 | 0.082 |
| Social apathy | 0.14 | 0.15 | 0.93 | 0.355 | 0.492 |  |  |
| Cognitive function symptoms | 0.27 | 0.16 | 1.67 | 0.103 | 0.240 |  |  |
| Depressed mood/suicidality | 0.13 | 0.17 | 0.81 | 0.422 | 0.492 |  |  |
| Physical anergia/slowness | 0.16 | 0.16 | 1.00 | 0.324 | 0.492 |  |  |
| **C. Cognitive effort cost, MDD, controlling for anxiety** |  |  |  |  |  |  |  |
| Anhedonia | -0.06 | 0.14 | -0.40 | 0.688 | 0.991 |  |  |
| Behavioral apathy | -0.06 | 0.14 | -0.40 | 0.689 | 0.991 |  |  |
| Social apathy | 0.01 | 0.13 | 0.07 | 0.947 | 0.991 |  |  |
| Cognitive function symptoms | 0.001 | 0.17 | 0.01 | 0.991 | 0.991 |  |  |
| Depressed mood/suicidality | 0.13 | 0.19 | 0.67 | 0.508 | 0.991 |  |  |
| Physical anergia/slowness | 0.06 | 0.16 | 0.39 | 0.696 | 0.991 |  |  |

#### Symptom specific relationships

We fitted regression models to estimate symptom domain relationships to effort costs while controlling for high-effort travel task performance, age, education, and BMI for physical effort (see Figure S.I. 10 for heatmap of symptoms and task correlations). Anxiety was related to *decreased* cognitive effort cost (Figure 2, Table 2), and this pattern was maintained in the current MDD group, across all participants (Table S.I. 9), and when controlling for medication use (Table S.I. 6). Given our inclusion of participants with comorbid anxiety disorders and prior literature relating anxiety to increased effortful model-based strategy, we tested whether statistically accounting for anxiety would reveal any other any other symptoms relationships to cognitive effort cost when and found no reliable relationships (Table 2).

We examined symptom associations with physical effort costs. Within the MDD group, anhedonia and behavioral apathy were associated with *increased* physical effort costs (Table 2). These effects were maintained (1) when controlling for medication (though not after FDR correction, Table S.I. 6), (2) in the current MDD group only (Table S.I. 9). In all participants, there was no reliable association with behavioral apathy, and the association with anhedonia was significant only before FDR correction. There was a significant difference in the correlation magnitudes of cognitive and physical effort cost with anxiety (z=-2.15, *p<*0.031), anhedonia (z=-2.71, *p<*0.007) and behavioral apathy (z=-2.69, *p<*0.007) within the MDD group.

### Additional task measures

#### Travel task performance

All the effort cost symptom analyses are over and above any effects related to travel task ability (i.e., task performance), which was controlled for. We examined diagnostic group differences and performance associations to symptoms directly (S.I. section 18). There were minimal diagnostic group differences in performance; however, the MDD group completed a lower percentage of keypresses in the high physical effort condition and responded faster on average on the cognitive task (Figure S.I. 11, Table S.I. 11). Neither cognitive nor physical performance was reliably related to overall depression. While anxiety symptoms were associated with cognitive effort costs, they were not associated with cognitive task performance (Figure S.I. 12, S.I. section 18). Anhedonia symptoms were related to a lower percentage of completed keypresses in the low (but not high) physical effort condition (*p<*0.010). Cognitive task performance did not predict cognitive effort cost, nor did physical performance predict physical effort cost, suggesting effort decisions and execution are dissociable in this task (Figure S.I. 12).

#### Overall exit threshold

We did not observe diagnostic group differences in overall exit thresholds in a model controlling for age and education (*p>*0.339). However, when medication was a covariate, medication use was associated with lower exit thresholds, and MDD group membership was associated with higher thresholds (Table S.I. 6). Overall exit thresholds were lower in participants with greater overall depression (Table 3, but not for other symptom domains), and this association was significant within the current depressed group (Figure 3, Table S.I. 12). The overall depression effect was maintained when controlling for medication (and additional effects were found for depressed mood/suicidality and physical anergia/slowing, Table S.I. 6). Overall, this pattern is consistent with theories of reduced subjective reward rate representation associated with depression (Huys et al., 2015).

**Table 3:** Symptom overall exit threshold regressions (MDD group only). Predicting individual differences in overall exit thresholds (log, from low effort conditions) by symptom severity, controlling for age and years of education (* indicates *p<*0.05, FDR correction within symptom models). Overall depression model *R*^2^=0.085 and adjusted *R*^2^=0.029. All variables were scaled as input to the regressions.

| Symptom | Estimate | SE | *t* | *p* | *p* *adjusted* |
| --- | --- | --- | --- | --- | --- |
| **Overall exit threshold, MDD** |  |  |  |  |  |
| Overall depression | -0.33 | 0.16 | -2.09 | 0.042* |  |
| Anhedonia | -0.26 | 0.14 | -1.80 | 0.078 | 0.109 |
| Anxiety | -0.34 | 0.14 | -2.43 | 0.019 | 0.066 |
| Behavioral apathy | -0.30 | 0.15 | -2.09 | 0.042 | 0.074 |
| Social apathy | -0.10 | 0.14 | -0.73 | 0.470 | 0.470 |
| Cognitive function symptoms | -0.21 | 0.15 | -1.44 | 0.157 | 0.184 |
| Depressed mood/suicidality | -0.40 | 0.15 | -2.68 | 0.010 | 0.066 |
| Physical anergia/slowness | -0.32 | 0.14 | -2.20 | 0.033 | 0.074 |

**Figure 3:**
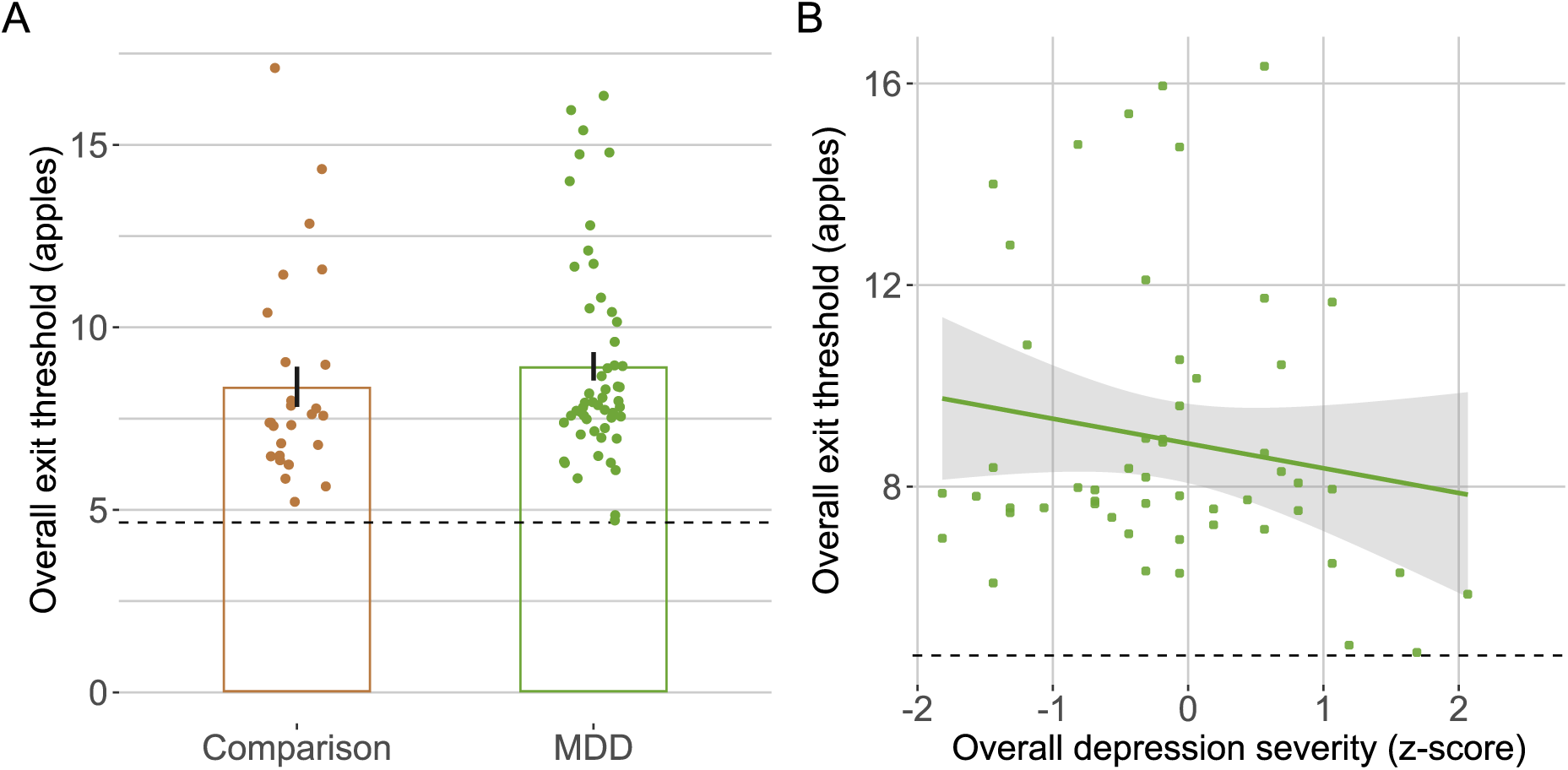
Relationship of individual MDD symptom domains with overall exit threshold (MDD group only). A: No diagnostic group differences (x-axis) in overall threshold (y-axis, apples, estimated from low effort conditions). Bar indicates group means, error bars indicate standard error of the mean, points indicate mean overall exit threshold per participant (i.e., random effects coefficients from linear regression model). B: Lower overall exit threshold (y-axes) were significantly related to overall depression severity (x-axes, Hamilton Depression Rating Scale Total z-score). Dashed line indicates best threshold policy, linear regression line for MDD group only.

## Conclusions

This cross-sectional study compared cognitive and physical effort-based decision-making using computational-model-derived parameters from the Effort Foraging Task (Bustamante et al., 2023), within-participant, in a heterogeneous group of participants with MDD and non-psychiatric comparisons. We found novel and important dissociable relations between symptom dimensions of MDD and cognitive versus physical effort. Our results corroborate several computational theories of depression and support breaking depression down into symptom domains and to examining components of effort- and value-based decision-making within-participants.

### Diagnostic group differences

We predicted MDD would be associated with increased effort avoidance for both cognitive (Ang et al., 2023; Hershenberg et al., 2016; Vinckier et al., 2022; Westbrook et al., 2022) and physical (Isabel M. Berwian et al., 2020; Cléry-Melin et al., 2011; Treadway et al., 2012; Vinckier et al., 2022; X.-H. Yang et al., 2014; Zou et al., 2020) effort. Contrary to our hypotheses, we did not observe significant group differences in effort costs. This aligns with null results in other studies of cognitive (Barch et al., 2023; Tran et al., 2021) and physical effort avoidance (Cathomas et al., 2021; Sherdell et al., 2012; Tran et al., 2021; Wang et al., 2022; X. Yang et al., 2021). There were minimal task *performance* differences, except the MDD group had faster N-Back reaction times and completed fewer keypresses in the high physical effort condition. Travel task performance and effort costs were not correlated in this sample, suggesting they are dissociable in this task.

### Symptom associations with effort costs

Greater overall depression severity in the MDD group was associated with greater willingness to exert cognitive effort, with no such relationship for physical effort. Although the comparison of correlations did not indicate specificity with respect to effort type. Anxiety symptom severity accounted for the cognitive effort cost association, whereas physical effort cost was not reliably related to anxiety. The comparison of correlations indicated effort type specificity. To our knowledge no effort-based decision-making studies have reported decreased cognitive effort avoidance associated with MDD, suggesting unaccounted anxiety variation might contribute to inconsistent findings.

The negative association between anxiety and cognitive effort cost is consistent with reports of increased model-based planning associated with anxiety in unselected samples (Gillan et al., 2016; Hunter et al., 2018). Increased cognitive effort exertion in anxiety might act as a compensatory mechanism to maintain performance amid threat-related attentional demands (Eysenck et al., 2007). Clinically, increased willingness to exert cognitive effort may contribute to anxiety symptoms such as rumination and worry through increased planning and replay (Bedder et al., 2023). Higher effort tasks might reduce anxious thoughts due to increased cognitive load (presumably via distraction). This is consistent with research showing reduced momentary anxiety during a high relative to low cognitive effort task (Vytal, Cornwell, Arkin, and Grillon, 2012).

Despite aiming to minimize demand characteristics, the social context of the experiment may have motivated anxious participants to exert effort (rather than expressing underlying preferences). An online study with the Effort Foraging Task found anxiety was associated with increased cognitive effort cost, opposite to present findings (Bustamante et al., 2023). However, the study was conducted in a large, unselected sample with self-reported symptoms, complicating translation to this clinical sample (another online study also did not report cognitive effort avoidance relationships to anxiety and depression, Patzelt, Kool, Millner, and Gershman, 2019).

Anhedonia and behavioral apathy were significantly associated with increased physical effort costs within the MDD group, when controlling for psychotropic medication, and the current MDD group, but not across all participants. The anhedonia association is consistent with some reports, (Sherdell et al., 2012; Tran et al., 2021; X.-H. Yang et al., 2014), but not others, (I. M. Berwian, Walter, Seifritz, and Huys, 2017; Cathomas et al., 2021; Cléry-Melin et al., 2011; Vinckier et al., 2022; Wang et al., 2022; X. Yang et al., 2021; Zou et al., 2020). Behavioral apathy has previously not been found to be reliably related to physical effort avoidance (Cathomas et al., 2021; Cléry-Melin et al., 2011; Vinckier et al., 2022). Willingness to exert cognitive effort was not related to anhedonia or behavioral apathy and comparison of correlations demonstrated differential relationships by effort type.

These findings support measuring both cognitive and physical effort decision-making function markers, which may inform heterogeneity or subtypes of MDD. Two lines of evidence suggest observed relationships between behavior and symptoms were driven by motivation rather than ability. First, all analyses relating effort costs to symptom severity accounted for ability (i.e., high effort performance) such that reported effects are over and above effects related to ability. Second, we found no reliable direct relationships between ability and MDD symptoms. The differential associations between anxiety versus anhedonia/behavioral apathy may suggest dissociable symptom dimensions caused by cognitive versus physical factors respectively (however, the present study was limited in teasing apart cognitive versus somatic forms of anxiety). Future studies can test whether anhedonia stems more from physical factors such as peripheral symptoms (e.g., fatigue), whereas anxiety is driven more by cognitive factors.

The Effort Foraging Task was developed to measure effort preferences more implicitly to increase validity. Therefore, methodological differences from previous (explicit) effort tasks may have contributed to the identification of symptom relationships, which has been mixed in other studies. On the other hand, implicit and explicit decisions may reflect unique effort-based decision-making aspects that differentially relate to MDD. It remains a question whether results from this task are more valid or if they tap into a different dimension of effort-based decision-making than the explicit task literature. If this were the case, it would enhance the novel contribution but also limit generalizability of the findings. Nevertheless, this work opens a new avenue for understanding how effort-based decision making relates to depression and other psychiatric symptoms, and how variations in task and modeling approaches affect such relationships.

### Subjective reward rate in depression

The MVT predicts that decisions to leave a patch reveal perceived environmental quality (i.e., opportunity cost of time), and this is supported by evidence in humans (e.g., lower thresholds under acute and chronic stress, in persons with Parkinson’s, in persons with opioid use disorder, associations with dopamine receptor availability, Constantino et al., 2017; Constantino and Daw, 2015; Ianni et al., 2023; Lenow et al., 2017; Raio et al., 2021). Following Huys et al. (2015), we hypothesized MDD would be associated with lower exit thresholds, reflecting reduced average reward expectations. Few studies have linked psychiatric symptoms to sensitivity to opportunity cost of time, though one found an association with self-reported apathy (Nair et al., 2023). Overall exit thresholds did not differ by group (though a group effect emerged when accounting for psychotropic medication use in an unexpected direction) but were decreased in MDD participants with greater overall depression severity (also when excluding remitted participants). This suggests a difference in value-based decision-making associated with the severity of MDD symptoms, such that environments may be subjectively represented as less rewarding, reducing goal-directed behavior and vigor (Huys et al., 2015; Niv et al., 2007).

### Limitations

These results leave open the question of whether observed symptom associations generalize to other psychiatric conditions or other effort-based decision measures (e.g., tasks, ecological momentary assessment). To determine if the association between anxiety and cognitive effort cost is specific to MDD, future studies could include participants with primary clinical anxiety disorders. Another limitation is that sample size of the remitted depressed group did not allow for comparison with other groups. Heterogeneity in psychotropic medication use is another limitation, given neurotransmitter effects on aspects of cognition measured in the task. However, key effects were robust to excluding remitted participants and controlling for psychotropic medication use. The cross-sectional design limits understanding causality between symptoms and task behavior. Longitudinal designs could distinguish state versus trait influences on cognitive control and effort-based decision making and their interaction with symptoms.

### Clinical implications

Ultimately insights from this research may inform interventions to increase willingness to exert effort for individuals experiencing challenges with goal-directed behavior due to psychiatric disability. Therapies that use cognitive restructuring to target physical effort perception might be effective for addressing anhedonia symptoms. Therapies for depression may target subjective reward rate, possibly through pharmacological dopamine manipulations (Niv et al., 2007). For applications to anxiety, the causal direction of the association with cognitive effort cost should be established. Does reduced cognitive effort cost cause anxiety symptoms, or does anxiety cause the pattern observed (i.e., benefits of distraction)? The tendency to exert cognitive effort could be leveraged as a strength in treating anxious depression (e.g., positive fantasizing, more cognitively effortful therapies, novel therapeutic applications using distraction, Besten, van Tol, van Rij, and van Vugt, 2023).

## Availability of Data and Materials

The analysis code and data will be openly available at the Open Science Framework (OSF) upon publication.

## Data Availability

The analysis code and data will be openly available at the Open Science Framework upon publication.

## Acknowledgements

Thanks to Nicole Antkiewicz, Valerie Lilley, Yongjjing Ren, and Daniel Oliver for help with data collection. Thanks to Sam Zorowitz for technical assistance with remote task administration. Thanks to Sean Devine for help with the Hierarchal Bayesian model.

## Financial support

Funding Statement: This work was supported by the New Jersey Alliance for Clinical and Translational Science (grant number UL1TR003017), the New Jersey Health Foundation (grant number NJHF#PC158-23), and the National Institutes of Health (LB, grant numbers T32MH065214, T32DA007261).

## Competing interests

The authors declare none.

## Ethical standards

The authors assert that all procedures contributing to this work comply with the ethical standards of the relevant national and institutional committees on human experimentation and with the Helsinki Declaration of 1975, as revised in 2008.

## 1 Study overview

The study was conducted at the Rutgers-Princeton Center for Computational Neuro-Psychiatry by trained clinical researchers. The study was approved by the Rutgers University Institutional Review Board. Participants were recruited in outpatient clinics at Rutgers University, as well as via Google Ads. We recruited participants with MDD confirmed using the Structured Clinical Interview for DSM-5 (SCID-5, First, 2015) and no co-occurring psychiatric conditions except for anxiety disorders, which were permitted. MDD participants in remission (“no significant symptoms during the past 2 months”) and in partial remission (“symptoms are present but full criteria are not met, for a period less than 2 months”) were permitted. We also recruited a demographically matched comparison group without any psychiatric diagnosis. The study was administered remotely via secure web-based software (Zoom, except for 5 MDD participants who came an in-person session) and participants were compensated $20 per hour and an Effort Foraging Task bonus of up to $10. On the first session a clinical interviewer administered the SCID-5 and participants completed self-report surveys. On the second session participants completed the Effort Foraging Task. In a third session participants completed tasks assessing reward sensitivity and cognitive control recruitment in response to efficacy manipulations, which will be reported separately.

### 1.1 Inclusion and exclusion criteria

The study inclusion criteria were 1) between the ages of 18-65, 2) has the capacity to provide informed consent, and 3) is fluent in English, 4) score of 6 or higher on the Wechsler Test of Adult Reading (equivalent to standardized score of 53 - 60 for ages 18-64, Wechsler, 2001), 5) (MDD only) Meets DSM-5 criteria for MDD as confirmed by the SCID-5, (MDD only) if the participant is treated with anti-depressant medication, they are on stable treatment with this medication (i.e. no change in medication type, or substantial change in dose, for at least 4 weeks prior to participating in the study). The study exclusion criteria were 1) history of traumatic brain injury or head injury, 2) Intellectual disability or pervasive developmental disorder, 3) neurological disease, 4) has met DSM-5 criteria for a substance-use disorder within the last 6 months per the SCID-5 (with the exception of nicotine dependence, which was permitted), 5) received electroconvulsive therapy within the last 8 weeks, 6) left-handedness (due to keyboard set up for the Effort Foraging Task), 7) (comparisons only) meets DSM-5 criteria for any psychiatric diagnosis as confirmed by the SCID-5, 8) (comparisons only) current use of any psychotropic medication.

### 1.2 Diagnostic group matching

We used Pearson’s Chi-squared test to compare categorical diagnostic group differences (gender, race, ethnicity, total household income, occupational status, relationship status, alcohol frequency, alcohol amount, caffeine amount, tobacco use), and Welch two sample unpaired t-test to compare continuous diagnostic group differences (age, childhood income, years of education for self and parents), using the chisq.test and t.test functions of the stats package in the R language, RCoreTeam, 2015). If diagnostic groups were not matched on a demographic variable, we included it as a covariate in our group difference and symptom relationships to task behavior analyses.

## 2 Power analysis

We conducted a power analysis with the current sample size (using the pwr package, Champely et al., 2020). For the symptom regressions, power analysis indicated we could detect a medium effect size with 80% power with the current sample of 52 MDD participants (*F* ^2^=0.254, Cohen’s *F* ^2^ ’medium’ effect size between 0.15 and 0.35, J. Cohen, 1992). For the diagnostic group differences, power analysis indicated we could detect a medium effect size with 80% power with the current sample of 52 MDD and 27 comparison participants (D=0.673, Cohen’s D ’medium’ effect size between 0.5 and 0.8, J. Cohen, 1992).

## 3 Major depressive episode criteria

Participants were assigned to the MDD diagnostic group based on the SCID-5, which considers the lifetime history for the MDD diagnosis, and symptoms in the past two months to establish whether a participant is 1) currently depressed, meeting criteria for a major depressive episode any time in the past month, 2) in partial remission, either experiencing some symptoms but not meeting full criteria for a major depressive episode, or there is a period lasting less than two months without significant symptoms, or 3) in full remission, during the past two months no significant symptoms. All participants in the MDD group have a lifetime history of depression, but varied in how many symptoms they experienced in the past week, as well as in the past two months. This dynamic variation in MDD symptom expression, if unaccounted for, may contribute to mixed results surrounding diagnostic group differences in effort-based decision making. This motivated us to focus on individual differences in symptom severity at the time of the study, rather than solely diagnostic group differences. The present study utilized the HAMD, which measures symptoms in the past week, along with the BPRS which measures symptoms in the past 2-3 days, as well as self-reports which asked about either the past few days, two weeks, or month. Therefore, currently depressed participants may have low scores on the HAMD (or BPRS) if they report fewer symptoms in the past week relative to the past month (during which they did meet full criteria for a major depressive episode). Low scores on the HAMD (or BPRS) would be expected for fully or partially remitted MDD participants, although scores could be higher for these participants if they are experiencing some symptoms but did not meet full criteria for a major depressive episode.

## 4 Clinician ratings and self reports

The Structured Clinical Interview for DSM-5 (SCID-5, First, 2015) was used to confirm assignment of MDD, co-morbid anxiety, and comparison groups, and that participants met study diagnostic inclusion and exclusion criteria. The clinical interviewer used responses in the SCID-5 to rate the severity of different symptoms in the past week via the the Hamilton Depression Rating Scale (HAMD, Hamilton, 1960) and to determine whether MDD participants were currently depressed, or in partial or full remission. This was followed by the semi-structured interview the Brief Psychiatric Rating Scale (Overall and Gorham, 1962) to assess current psychiatric-symptom severity. Next participants completed self-report surveys to measure; cognitive function symptoms and physical anergia with the Massachusetts General Hospital Cognitive and Physical Functioning Questionnaire (Fava, Iosifescu, Pedrelli, and Baer, 2009), depression symptoms with the Patient Heath Questionnaire-9 (Kroenke, Spitzer, and Williams, 2001), anxiety symptoms with the Generalized Anxiety Disorder-7 (Spitzer, Kroenke, Williams, and Löwe, 2006), anhedonia symptoms with the Snaith–Hamilton Pleasure Scale (Nakonezny, Carmody, Morris, Kurian, and Trivedi, 2010), behavioral, emotional, and social apathy with the Apathy Motivation Index (Ang, Lockwood, Apps, Muhammed, and Husain, 2017). We also measured trait executive function with the Adult Temperament Questionnaire Effortful Control subscale (Evans and Rothbart, 2007), and trait cognitive control seeking with the Need for Cognition scale (Cacioppo, Petty, and Kao, 1984).

## 5 Symptom confirmatory factor analysis

We performed confirmatory factor analysis and assigned items from clinician ratings and self-report into the following domains (using measures listed in Table S.I. 1, exact items in Table S.I. 3): anhedonia, appetite symptoms, anxiety, behavioral apathy, cognitive function symptoms, depressed mood, effortful control (trait), emotional apathy, need for cognition (trait), physical anergia/slowing, social apathy. Assigned items were z-scored and averaged to compute a symptom score in (1) the MDD group only and (2) all participants. Confirmatory factor scores for comparison participants did not include ratings from clinician measures (i.e., HAMD, and BPRS) and instead were the average z-score of all self-report items in a factor.

We tested whether diagnostic groups significantly differed in symptom intensity and cognitive control trait measures using t-tests (Table S.I. 2). For left skewed distributions (comparisons clustered on very low symptom scores for anxiety, cognitive function symptoms, depressed mood, and physical anergia/slowing) we confirmed the diagnostic group differences results were maintained using the non-parametric Wilcoxon rank sum test with continuity correction (wilcox.test function of the stats package in the R language, RCoreTeam, 2015).

## 6 Effort Foraging Task counterbalancing

The order of cognitive and physical effort variants was counterbalanced across participants. Within blocks of an effort type, each effort level was tested once during the first half and once during the second half. Given that constraint, the effort level was fully counterbalanced, resulting in eight possible orders. Participants were assigned a block order using latin squares within each diagnostic group.

## 7 Task instructions and training

“Welcome to the experiment! Thank you for participating. This experiment will require you to press buttons on your keyboard repeatedly, applying varying amounts of physical effort. If you have any history of any sort of hand injury or pain with typing (e.g., which could make either fast button pressing or stretching your hand uncomfortable) please do not complete this task. You must wait a minimum of 5 seconds before you are able to progress to a new slide of instructions. You will know you can click to a new slide when the “Next” button changes.”

“Welcome to the Apples Game! For your completion of this task, you will receive a potential bonus between $0 and $5. Please read the instructions carefully. There will be a quiz at the end of these instructions to check your understanding. In this game, you will make choices that earn you money. Imagine you are a farmer, and you are harvesting apples from trees in your multiple orchards. On every trial within an orchard, you will see a tree: To HARVEST the tree, press the down arrow key with your right hand. Do this when the circle below the tree is white. When you harvest the tree it gives you apples. These apples are worth real money that you will earn on top of the money for participating in this study. Now, try harvesting the tree three times in a row. Press the HARVEST key to collect apples from the tree. [Press “Next” to practice using the HARVEST key 3x]”

“The more times you harvest a tree, the fewer apples it gives you! On any trial, instead of accepting the number of apples the tree is giving you, you have the option to TRAVEL to a new tree. To TRAVEL to a new tree press the right arrow key with your right hand. Do this when the circle below the tree is white. Now, harvest the tree once and travel from one tree to another. Do this three times. Press the TRAVEL key to move to a new tree. [Press “Next” to practice using the TRAVEL key 3x]”

“Different trees give you different number of apples at the start. Exactly how many apples a tree starts with changes from tree to tree. The starting number of apples for a tree is NOT RELATED to how the tree looks or which orchard you are in. HARVESTING takes some time but earns you apples. TRAVELING takes longer, and you cannot harvest apples during traveling. But it brings you to a new tree with a full supply. You have to decide how to spend your limited time in an orchard – harvesting or traveling. You have to HARVEST each new tree once before traveling away from it. If you take too long to make a choice you will miss a turn and see this message. The more turns you miss, the less time you have to harvest apples, and you will earn less apples. That is most of what you have to know to be a great farmer. Let’s go through a short practice orchard in the Apples Game.”

Then participants completed a 1.5-minute orchard with no travel task. Then the learned one of the effortful travel tasks (order of effort types counterbalanced).

### 7.1 Foraging task training

In the task began with training the travel task for the first effort cost variant for a particular participant (this could be the cognitive or physical effort task). Next came instructions for the foraging task in general (without mentioning the effortful travel requirement), and participants completed a practice block (90 seconds) of the foraging task with no travel task. Then participants were instructed that they would have to complete the effortful travel task when traveling, and they completed two practice blocks (one per effort level, 90 seconds each). Then, participants completed the main foraging task for the first travel task type (4 blocks, 7 minutes per block, with self-paced breaks between blocks). After completing all the blocks of the first travel task, participants began training on the second travel task. They were instructed that they would continue to play the foraging task, but the travel task had changed. They practiced the foraging task with the second travel task type (one practice block per effort level, 90 seconds each). Finally, they completed the main foraging task for the second travel task type (4 blocks, 7 minutes per block).

### 7.2 Rapid key-pressing task training

In Experiment 1 key-press training began with a calibration phase (three rounds) to determine the maximum number of presses participants were able to complete in the travel time (7.5 seconds of effort task time). A counter was displayed on the center of the screen showing how many presses a participant had made. The instructions suggested participants were being compared to others, and encouraged them to press as fast as possible, each round they were encouraged to press faster than they had the previous rounds. Then we used each participant’s mean number of presses across rounds as their ‘maximum number’. We enforced a minimum ‘maximum number’ value of 20 presses. The Larger Number of Presses condition tasked participants with completing 100% of their maximum, and the Smaller Number of Presses condition tasked participants with completing 50% of their maximum. Participants were told that there was a larger and smaller number, but not what that number was or how it was determined. Then participants practiced a single effort level. Effort level order was counterbalanced. Practice for an effort level began with a single mini-block the duration of the foraging travel time. Then participants had to complete 5 mini-blocks reaching the required number of presses to move on. This was meant to establish the expectation that participants would perform well on the travel task, even though there were no incentives or punishments associated with travel task performance during the foraging task.

#### 7.2.1 N-Back working memory task

The N-Back task was performed as part of foraging task during travel between trees. In the N-Back task letters are displayed on screen in a sequence. Participants judged whether the stimulus that is currently on the screen matches the stimulus they saw a number of screens back (N-Back). On every trial, participants responded whether the letter was a match (“s” key) or non-match (“d” key) to the letter on the previous screen (1-Back case) or three screens before (3-Back case). A trial began with a fixation cross (for 250 milliseconds) followed by the letter on screen (for 500 milliseconds) followed by a blank screen (for 950 milliseconds, total trial duration = 1.7 seconds). During the travel period, 10 letters were presented, of which, 2 or 3 were targets (letter matches letter N-Back) and 2 or 3 were lures (matches current letter but not in position N-Back). The number of targets and lures were selected randomly each time an N-Back stimulus sequence was generated. We only used consonants to prevent participants from using mnemonics (letters were: ‘B’, ‘C’, ‘D’, ‘F’, ‘G’, ‘H’, ‘J’, ‘K’, ‘M’, ‘N’, ‘P’, ‘Q’, ‘R’, ‘S’, ‘T’, ‘V’, ‘W’, ‘X’, ‘Y’, ‘Z’), and half of the letters were presented in upper case and the other half lower case to prevent participants using iconic memory (J. D. Cohen et al., 1994).

#### 7.2.2 N-Back working memory task training

We trained the N-Back task extensively to try to bring participants to highest possible levels of performance and minimize automaticity differences (in which some participants would have more experience with the N-Back or similar tasks, making the task less effortful for them compared to someone with little experience). Participants had to reach a performance criterion to move on from training. After being instructed on the task participants began practice for one of the effort levels (counterbalanced). First, they completed two extended blocks (50 trials with a self-paced break up to 45 seconds between) with feedback about error type (types of feedback: “non-match”, “missed match”, “no response”, displayed in red font for 800 ms after the trial). Then they performed one extended block without any feedback (50 trials).

We tasked participants with completing a set number of mini-blocks with high accuracy to begin the foraging task. We did so to establish the expectation that participants had to exert effort when they chose to travel while foraging. A mini-block was classified as successful when the participant saw no error feedback (large black dot), after which they were told they were moving on to the next mini-block. The error feedback was displayed when participants made two consecutive errors (including omission errors). If they did see one or more error feedback symbols, they had to repeat that mini-block. They had to successfully complete 8 mini-blocks of the 1-Back task, and 12 mini-blocks of the 3-Back task. This training also ensured that participants could adequately perform the task. Participants had self-paced breaks in between mini-blocks (up to 60 seconds).

## 8 Foraging behavior analysis exclusions

Of the 60 MDD and 27 comparison participants, 1 MDD participant did not complete the Effort Foraging Task due to technical difficulties with their keyboard. All other participants completed the task, however technical difficulties with the experiment server caused 4 missing data files from the MDD group. We followed a subset of exclusions validated in Bustamante et al. (2023) that most interfere with estimating effort costs. First, participants were excluded if they had very few exit trials within an effort type, making their data under-powered for estimating exit thresholds, and overly deterministic for logistic regression, which are the basis of the effort cost measures (2*SD below the mean, *<*8.82 trials). As a result 1 MDD participant was excluded from analysis for the whole task (1 exit in high effort physical and 3 exits in high effort cognitive condition) and 1 MDD participant was excluded from the cognitive effort analyses (2 exits for the cognitive high effort condition). Second, participants were excluded from the task if they missed the response deadline on many foraging trials (2*SD above the mean, *>*15.05%, 1 MDD participant excluded who missed 49.5% of trials) which may reflect low engagement with the task or challenges meeting the response deadline. Ultimately, this affects the interpretability of MVT estimates (e.g., experienced harvest time longer than for other participants, fewer apples per second). The final sample included in behavioral analyses was 52 MDD participants (53 MDD participants in the physical effort condition) and 27 comparison participants.

## 9 MVT model additional methods

Because we are investigating individual differences in effort costs at the condition level, used a factorial model in which the MVT threshold is taken as fixed per-condition, determined by the overall rewards and delays in each condition and a per-condition effort-cost parameter. Thus, the model omits trial-by-trial learning of the threshold, and instead formally absorbs any such variation into the softmax choice stochasticity. We believe this simplification is warranted because the condition-wise effort costs of interest aggregate over per-trial threshold variability, and because we encouraged asymptotic behavior through pre-training and using a stable foraging environment throughout.

There were five parameters in the model, the inverse temperature (*β*, which controls the noise of the softmax choice function, with lower values indicating more noisy effects of rewards and thresholds on choices), the cognitive low (*c*_cog low effort_) and high effort costs (*c*_cog high effort_), and the physical low (*c*_phys low effort_ and high effort costs (*c*_phys high effort_). The model included a full covariance matrix of the parameters (5-by-5 matrix) which consists of a correlation matrix and a scale (standard deviation) matrix. Parameters were drawn from a multi-variate Gaussian distribution. We used the covariance matrix to estimate the correlation between individual differences in high cognitive and physical effort costs. Model priors were centered at zero and variances were selected to accommodate the magnitude of group-level posterior distributions from the original Effort Foraging Task study (Experiment 2, Bustamante et al., 2023). The prior distributions for group-level effects were *c*_low effort_ *∼ N* (0, 25), *c*_high effort_ *∼ N* (0, 15), *β ∼ N* (0, 1). The prior on random effects variances were *c*_low effort_ *∼ N* (0, 25), *c*_high effort_ *∼ N* (0, 15), *β ∼ N* (0, 1). The prior on the correlation matrix was unbiased as to the presence or absence of a correlation (LKJ Correlation Distribution prior=1, Lewandowski, Kurowicka, and Joe, 2009). Individual participant parameters and their group-level distributions were estimated using Markov Chain Monte Carlo sampling, implemented in Stan with the CmdStanR package (4,000 samples, 2,000 warm-up samples, across four chains, Stan Development Team, Stan, 2021). Convergence was assessed by visually inspecting model traces, and ensuring the R^^^ convergence diagnostic statistic was below 1.1. We also simulated the MVT model to estimate the best exit threshold with respect to reward and time given the foraging environment parameters. To test for diagnostic group (*g*) differences, we fitted a Hierarchical Bayesian MVT model in which each of the 5 group-level parameters (*p*) had a diagnostic group effect (*β_g,p_*) added to it. For a participant (*i*) in diagnostic group (*g_i_*) each participant-level parameter *p_i_*was the sum of the group-level parameter *p* and the diagnostic group effect (*p_i_*= *p* + *β_g,p_ ∗ g_i_*, where *g_MDD_, g_Comparison_ ∈* 0.5*, −*0.5). The diagnostic group effect parameters (*β_g,p_*) for low and high effort travel costs had a prior distribution *N* (0, 5), and inverse temperature had a prior distribution *N* (0, 0.5), values greater than zero indicate higher effort costs in the MDD relative to comparison group. To confirm the remitted and partial remitted participants did not change the results, we also used this model to test for group differences of the comparison group to the currently depressed subset of the MDD group.

## 10 MVT model evaluation

We used several methods to evaluate the MVT model fit. We inspected trace plots to ensure mixing between chains, and the R^^^ convergence diagnostic was below 1.1 for all parameters (using the rhat and mcmc trace functions from the bayesplot package in R, Gabry et al., 2024). We conducted a posterior predictive check to confirm the fitted model captured foraging decisions. For each of 8,000 MCMC samples, for all trials across the entire dataset, the model sampled from the posterior predictive distribution from a Bernoulli distribution, generating a set of harvest (1) or exit (0) choices (using the bernoulli logit rng function in Stan, Team, 2021). We examined the correspondence of the posterior predictive samples to the empirical data. We tested whether the empirical probability of stay choices (across all participants and all trials in the dataset) fell within the posterior predictive distribution (i.e., the simulated probability of stay choices across all trials for each MCMC sample). To do so, we computed the distance from the median simulated probability of staying for every MCMC sample, as well as the distance from the empirical data. We tested the probability that the distance of the empirical data to the simulated median was larger than the distances of the simulated data. Additionally, we visually compared the overall exit threshold, as well as the change in exit threshold by diagnostic group, in simulated versus empirical data. Similarly, we visually compared the probability of exiting the patch across expected reward levels, as well as the change in the probability of exiting the patch by effort level (high - low effort conditions) across expected reward levels.

We assessed the log posterior likelihood per participant and used an unpaired t-test to compare goodness of fit for the model between the diagnostic groups. To do so, we computed the sum of log likelihoods within each participant, for each MCMC sample (using the bernoulli logit lpmf function in Stan, Team, 2021). Then we aggregated across MCMC samples by exponentiating these values, summing them, and log-transforming them, resulting in one log posterior likelihood value per participant (reflecting the logarithm of the posterior probability of the model parameters given the observed data for a participant). A lack of diagnostic group difference in this metric would suggest comparable goodness of fit.

## 11 Fatigue effects group differences methods

To measure fatigue we used linear mixed-effects regression predicting the model-agnostic measure of expected reward (log(apples)) by the effort level interacted with exit number within a round (starting at one to the total number of exits for a round) separately for cognitive and physical effort. Random effects terms were the effort level and exit number within a round, but without the interaction term due to convergence issues. We reasoned that fatigue should increase each time the travel task was completed. Because there were self-paced breaks between rounds we did not look at fatigue across rounds. Following this simpler model, we added the diagnostic group as a 3-way interaction with fatigue and effort level.

## 12 Simulation to find best threshold

We considered the best threshold found by simulation in Bustamante et al. (2023) for Experiment 2. To repeat the methods we used in that study, we simulated the best foraging threshold by creating a foraging environment with an agent with a fixed exit threshold and observing the resulting reward rate. We used a policy iteration algorithm to find the maximal reward rate for a given foraging environment. The foraging environment was defined by the following parameters from our experiments; the harvest time (2 seconds), travel time (8.33 seconds), the distribution of initial rewards to a tree *N* (15, 1) distribution of the decay function (beta distribution, *β*(14.90873, 2.033008)). We assumed the agent knew the mean depletion rate (0.88 multiplied by the previous reward) and used this value to predict the expected reward on the current trial. If the predicted reward was less than or equal to the agent’s threshold it exited the patch *R_e_ ≤ ρ*, otherwise it harvested the patch which yielded reward. We simulated 840 ‘seconds’ of foraging time for all experiments (though the result should be robust to duration). The simulation outputs were the ‘best threshold’ (threshold that yielded the highest reward rate, results vary slightly by simulation run), the resulting ‘best reward rate’, as well as the mean and standard deviation number of harvests to reach that exit threshold.

The agents’ threshold parameter was initialized at 4 apples. For an iteration i, the threshold was set as the mean reward rate observed in iteration i-1, this allowed the threshold to gradually improve in terms of reward rate between iterations. The simulation stopped and the best threshold was determined based on the stopping threshold of a 0.001 apple per second improvement in reward rate on iteration i compared iteration i-1 (with a maximum of 200 iterations).

## 13 Additional task measures methods

### 13.1 Task ability

Using a series of regression models, we tested whether diagnostic groups differed on effortful travel task performance. For the cognitive (N-Back) task we tested for differences in accuracy, reaction time, and missed trials. Using linear regression, we predicted N-Back accuracy (D’) by N-Back level interacted with diagnostic group. We used logistic mixed-effects regression to predict N-Back reaction times (log transformed) across all trials by a 4-way interaction between N-Back level, correct or incorrect response, target or non-target trial, and diagnostic group, controlling for age. We used logistic mixed-effects regression to predict the percent of missed N-Back trials by diagnostic group.

For the physical effort (rapid key-pressing) task we compared the groups on the required number of keypresses (determined during calibration) and the percent of keypresses completed during travel. Using linear regression, we predicted required keypresses by diagnostic group controlling for age. In a linear mixed effects regression, we predicted the percent of completed keypresses per travel interval by the effort level (smaller or larger number of presses) interacted with diagnostic group, controlling for age and BMI.

We tested whether cognitive and physical effort costs were dissociable from task ability (i.e., performance). Using data from all participants, in the first model we predicted cognitive effort cost by 1-Back and 3-Back D’, controlling for age. In the second model we predicted cognitive effort cost by the change in D’ from 1-Back to 3-Back (which in line with effort cost as a change score from low to high effort). In the third model we predicted physical effort cost by the percent of key presses completed in the larger number of presses, and the smaller number of presses condition, controlling for age and BMI.

### 13.2 Overall exit threshold

Overall exit threshold individual differences were estimated from a linear mixed effects regression model that predicted exit thresholds (log apples) in low effort orchards (which were least confounded by effects of effort) with effort type as a fixed effect, and random intercepts per participant. To test for diagnostic group differences, we added diagnostic group to the regression. To test for relationships with depression symptoms in the MDD group only, we ran a series of linear regressions predicting overall exit threshold by i) overall depression, and ii) each of the symptom domains separately (7 tests), controlling for age. We corrected for multiple comparisons across symptoms (FDR, 7 tests). We repeated these analyses zooming in to the currently depressed MDD group only and zooming out to all participants.

## 14 Relationship between cognitive and physical effort costs

Previously, we found a significant positive correlation between cognitive and physical effort costs in a large online study (Experiment 1 (MSIT) of Bustamante et al. (2023), N=537, mean correlation=0.566, 95% HDI=0.355, 0.766). In a smaller undergraduate sample (Experiment 2 (N-Back) of Bustamante et al. (2023), N=81) there was no conclusive evidence for or against the correlation, as the highest density interval (HDI) was wide (mean correlation=0.048, 95% HDI=-0.369, 0.462). The present study uses the same N-Back version of the task as Experiment 2 of the original study with a comparable sample size (N=80 participants in total) and yielded a similarly wide posterior distribution (mean correlation=0.0532, 95% HDI=-0.240, 0.345, pd=0.365, Table S.I. 5). In both cases the credible interval overlapped with the posterior distribution from Experiment 1, and it may simply be the sample size is underpowered to detect the presence or absence of a correlation. Beyond this, there are several other differences between these task versions, MSIT involves interference control, whereas N-Back involves working memory. Furthermore, the N-Back version is longer in duration (56 versus 32 minutes of main task time), the longer travel time (8.33 seconds versus 20 seconds) requiring more sustained effort. More research is needed to understand under which conditions cognitive and physical effort-based decision making are connected versus dissociated.

Ours and previous research on the relationship between individual differences in cognitive and physical effort decision making have found moderate correlations (e.g., correlation=0.43 in Lopez-Gamundi & Wardle, correlation=0.35 in Tran et al., 2020). This unshared variance between the effort domains leaves open the possibility of decoupling of their relationship to specific psychiatric symptoms. Indeed in Experiment 1 of Bustamante et al. 2023 (using the MSIT to elicit cognitive effort), we found cognitive, but not physical effort cost loaded strongly onto the dimension predictive of symptoms in a CCA (Dimension 1). Here, we demonstrated differential relationships of cognitive and physical effort cost to symptoms by conducting a comparison of correlations (Meng, Rosenthal, and Rubin’s z statistic, Meng, Rosenthal, and Rubin, 1992).

## 15 Diagnostic group differences in effort-seeking

We used Pearson’s Chi-squared tests with Yates’ continuity correction to test whether the proportion of participants with negative effort costs differed by diagnostic group. For cognitive effort, 22.2% of comparison participants and 20% of MDD participants had a negative effort cost and there was no significant difference between groups (chi-square*<*0.001, *df* =1, *p*=1). For physical effort 29.6% of comparison participants and 28.3% of MDD participants had a negative effort cost and there was no significant difference between groups (chi-square=0.0, *df* =1, *p*=1).

## 16 Model-agnostic sensitivity to effort manipulation

The model-agnostic measure, change in exit threshold, showed that on average exit thresholds were lower in the cognitive and physical, high relative to low effort conditions (3-Back - 1- Back estimate=-0.092 log(apples), *SE*=0.022, *df* =74.24, *t*=-4.212, *p<*0.001; Larger – Smaller Number of Presses: -0.056 log(apples), *SE*=0.0249, df = 76.83, *t*-2.233, *p<*0.0285). There was no reliable interaction between diagnostic group and change in exit threshold for cognitive (*t*=0.991, *df* =73.78, *p>*0.325) nor physical effort (*t*=0.673, *df* =76.88, *p>*0.503). The MVT model group-level posterior parameters indicated high effort cost is greater than zero for both effort types (Table S.I. 5). There was considerable individual variation in willingness to exert effort, signaling differences in perceived effort costs (see Figure 1).

## 17 Fatigue effects group differences results

For both cognitive (*t*=-2.437, *p<*0.016) and physical effort (*t*=-2.617, *p<*0.010) we found a main effect of trial number within a round on model-agnostic exit thresholds, suggesting that overall thresholds may have been impacted by fatigue. However, there were no interactions with effort level, suggesting this process was not differentially affected in high or low effort orchards (cognitive effort, *p>*0.085, physical effort, *p>*0.024). Given that the effort costs depend on the difference between conditions, if they are comparably affected by fatigue, this would be unlikely to explain the effort cost specific effects. However, there is a limitation of this measure, which is that as participants get close to the end of a round they might suspect it is not worth traveling if the block will timeout before they get to the next orchard. This would be consistent with the effect of trial number observed, which could also be a combination of these factors.

Next we examined the three-way interaction between effort level, exit trial number, and diagnostic group. There were no diagnostic group differences in the effects of exit trial number (two-way interaction, cognitive effort, *p>*0.901, physical effort, *p>*0.945), nor in the three-way interaction (cognitive effort, *p>*0.680, physical effort, *p>*0.105). Therefore, fatigue or time in block related effects are unlikely to explain the lack of group-level differences in effort costs.

## 18 Additional task measures results

Across all the travel task measures tested we found few reliable diagnostic group differences (see Figure S.I. 11 and Table S.I. 11), including no difference in missed N-Back trials, cognitive task accuracy (D’), nor required keypresses determined in the calibration phase. We found the MDD group responded faster on average on the cognitive (N-Back) task (Figure S.I. 11). We found a significant effect of diagnostic group on percent of completed presses, in which the MDD group completed a larger percent of presses across conditions, but a significant diagnostic group by effort level interaction, in which the MDD group completed fewer presses. To decompose this effect, we ran the same regression separately for each effort level. There was no reliable group effect on the percent of smaller number of presses completed, but the MDD group completed a lower percent of required keypresses in the larger press condition.

We found that cognitive and physical effort costs were dissociable from task performance in the Effort Foraging Task, this may suggest a disconnect between effort selection and effort execution (as suggested in, O’Reilly, Hazy, Mollick, Mackie, and Herd, 2014). There was no reliable association between cognitive effort cost and cognitive task performance (Figure S.I. 12, model 1: 3-Back D’, *p>*0.47, 1-Back D’, *p>*0.85, all participants, model 2: change D’, *p>*0.75). Likewise, there was no reliable relationship between physical effort cost and the percent of key presses completed (model 3: larger number of presses, *p>*0.83, smaller number of presses, *p>*0.31, consistent with that was found in Culbreth et al., 2023).

Neither cognitive nor physical performance was reliably related to overall depression (predict Hamilton Depression Rating Scale Total controlling for age by 3-Back D’, *p>*0.16, 1-Back D’, *p>*0.86). While anxiety symptoms were associated with cognitive effort costs, they were not associated with cognitive task performance (MDD group predict by anxiety symptoms controlling for age by 3-Back D’, *p>*0.21, 1-Back D’, *p>*0.79). On the other hand, anhedonia symptoms were related to a lower percentage of completed keypresses in the low (*t*=2.68, *p<*0.010), but not high (*p>*0.311) physical effort condition.

Overall depression was not related to cognitive task performance (Figure S.I. 12, predict HAMD total controlling for age by 3-Back D’, *p>*0.16, 1-Back D’, *p>*0.86) nor for physical task performance (predict HAMD total controlling for age and BMI by Larger number of presses, *p>*0.073, Smaller number of presses, *p>*0.370). While anxiety symptoms were associated with cognitive effort costs, they were not associated with cognitive task performance (MDD group predict by anxiety symptoms by 3-Back D’, controlling for age, *p>*0.21, and 1-Back D’, *p>*0.79). Anhedonia symptoms were related to the percent of smaller number of presses completed (*t*=2.70, *p<*0.010) but not to the percent of larger number of presses (*p>*0.318). Behavioral apathy symptoms were related to the percent of smaller number of presses completed (*p>*0.200) but not to the percent of larger number of presses (*p>*0.240).

---

**Figure S.I. 1:**
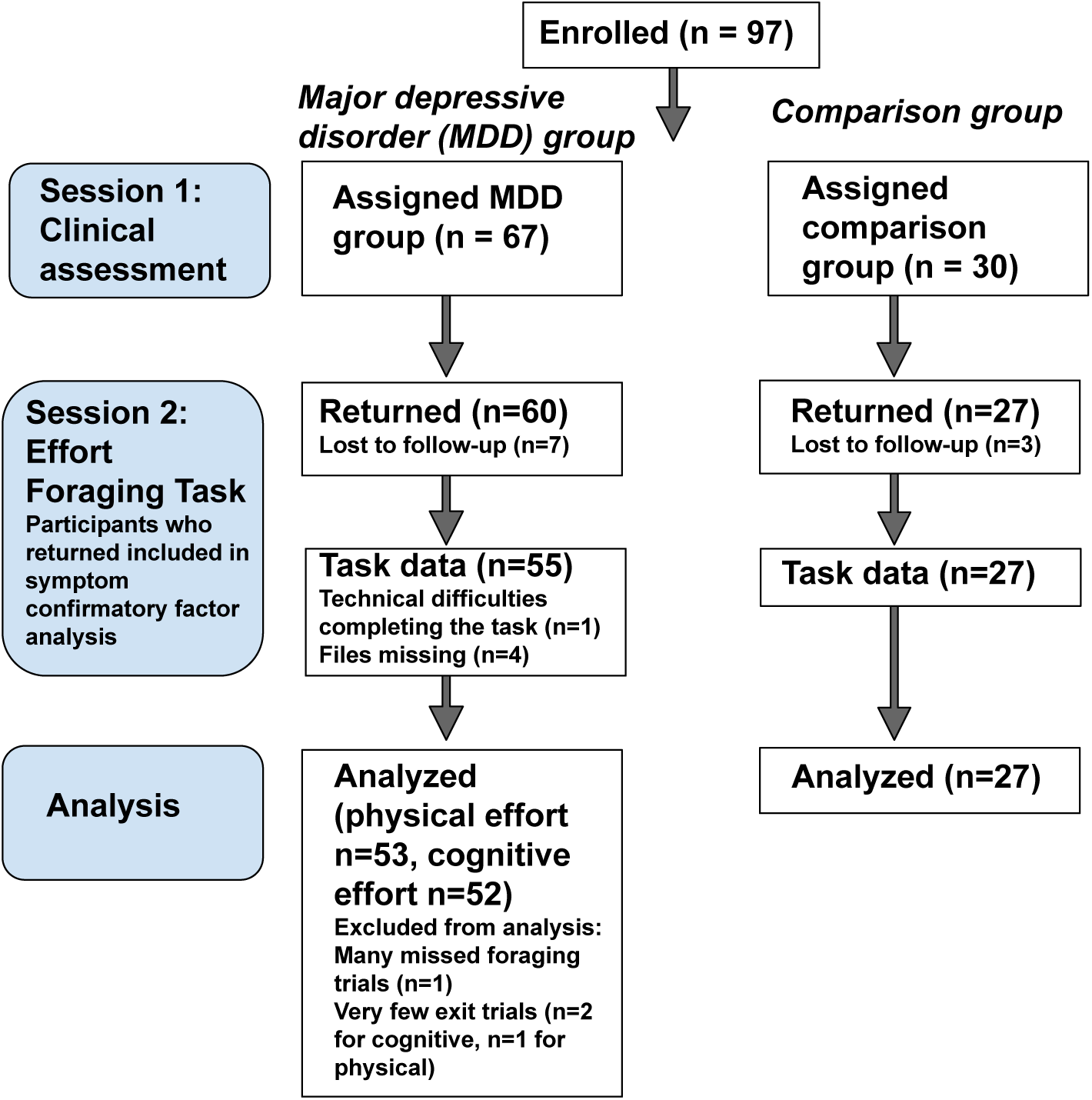
Inclusion and exclusion diagram. First column indicates experimental phase, session 1 clinical assessment, follow-up session 2 task measure, and number of participants analyzed. Second column indicates MDD group, third column indicates comparison group.

**Figure S.I. 2:**
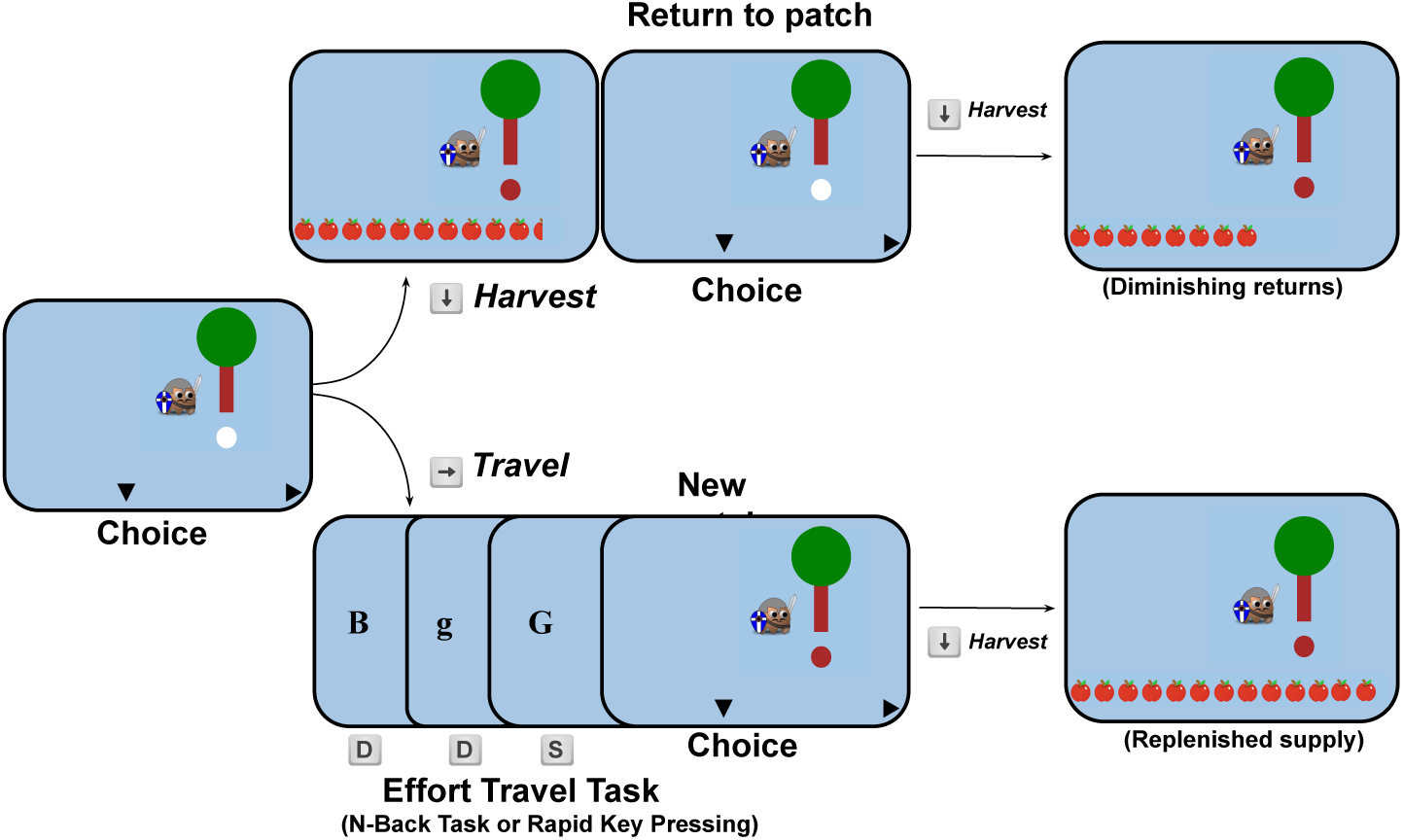
Effort Foraging Task Diagram. On each trial participants chose to harvest a virtual patch (apple tree) using the down arrow key, or travel to a new patch. Harvesting a patch yielded diminishing returns, whereas traveling to a new patch cost time and effort. Travel tasks were either the 1-Back or 3-Back levels of the N-Back task, or a smaller or larger number of rapid keypresses. Adapted from Figure 1 of Bustamante et al., 2023.

**Figure S.I. 3:**
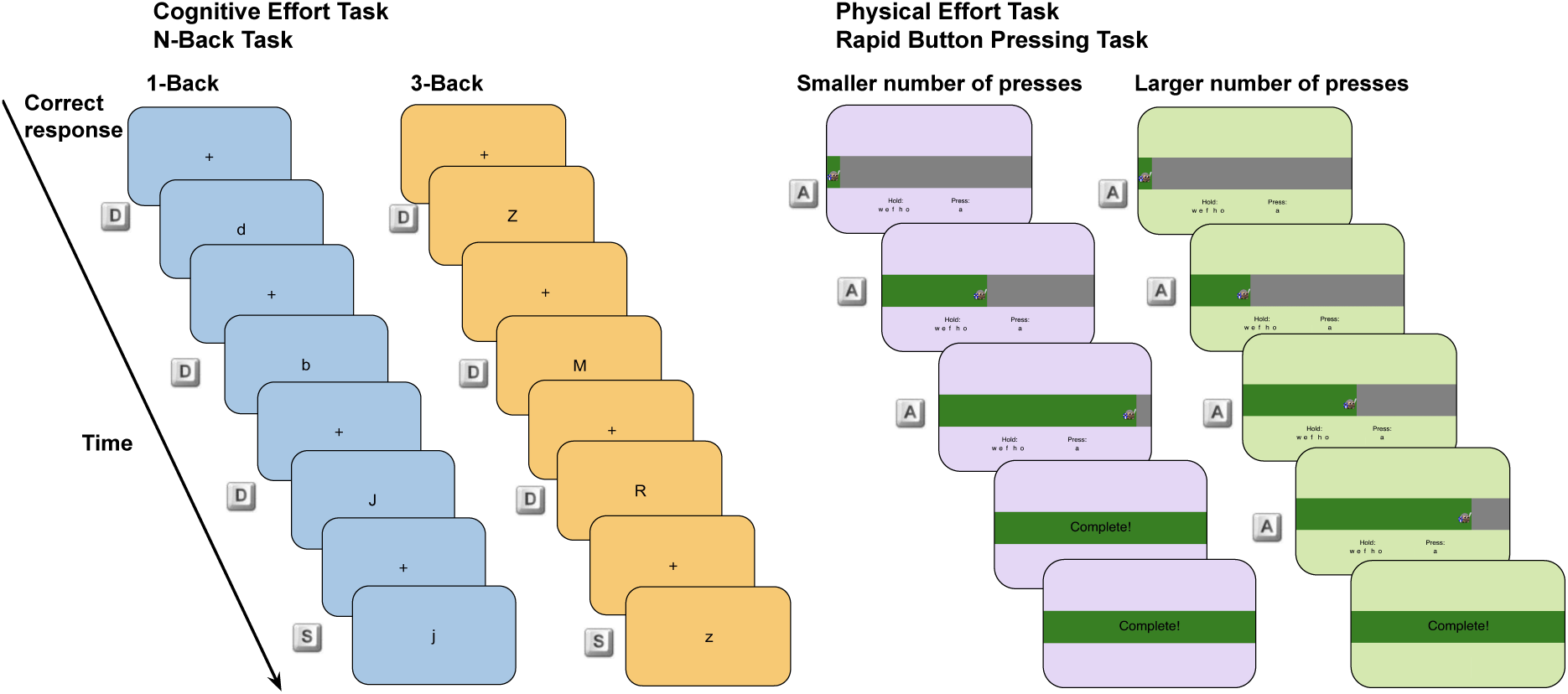
Travel task methods. Left panel: cognitive effort, N-Back working memory task. Participants responded whether the letter on the screen was the same (‘s’ key) or different (‘d’ key). The background color differed for the high effort (3-Back, orange) and low effort (1-Back, blue) conditions. Key icons next to each screen indicate the correct response. Right panel: physical effort, rapid key-pressing task. Participants rapidly pressed the ‘a’ key while holding down the ‘w’, ‘e’, ‘f’ (left hand), ‘h’ and ‘o’ keys (right hand). Pressing the ‘a’ key moved the avatar rightwards and filled up the grey horizontal bar with green. When participants reached the goal number of presses ‘Complete!’ appeared in the horizontal bar and participants waited for the remainder of the travel time. The background color differed for the high effort (smaller presses, 50% of maximum, purple) and low effort (larger presses, 100% of maximum, green) conditions. Adapted from Figure 2 of Bustamante et al., 2023.

**Figure S.I. 4:**
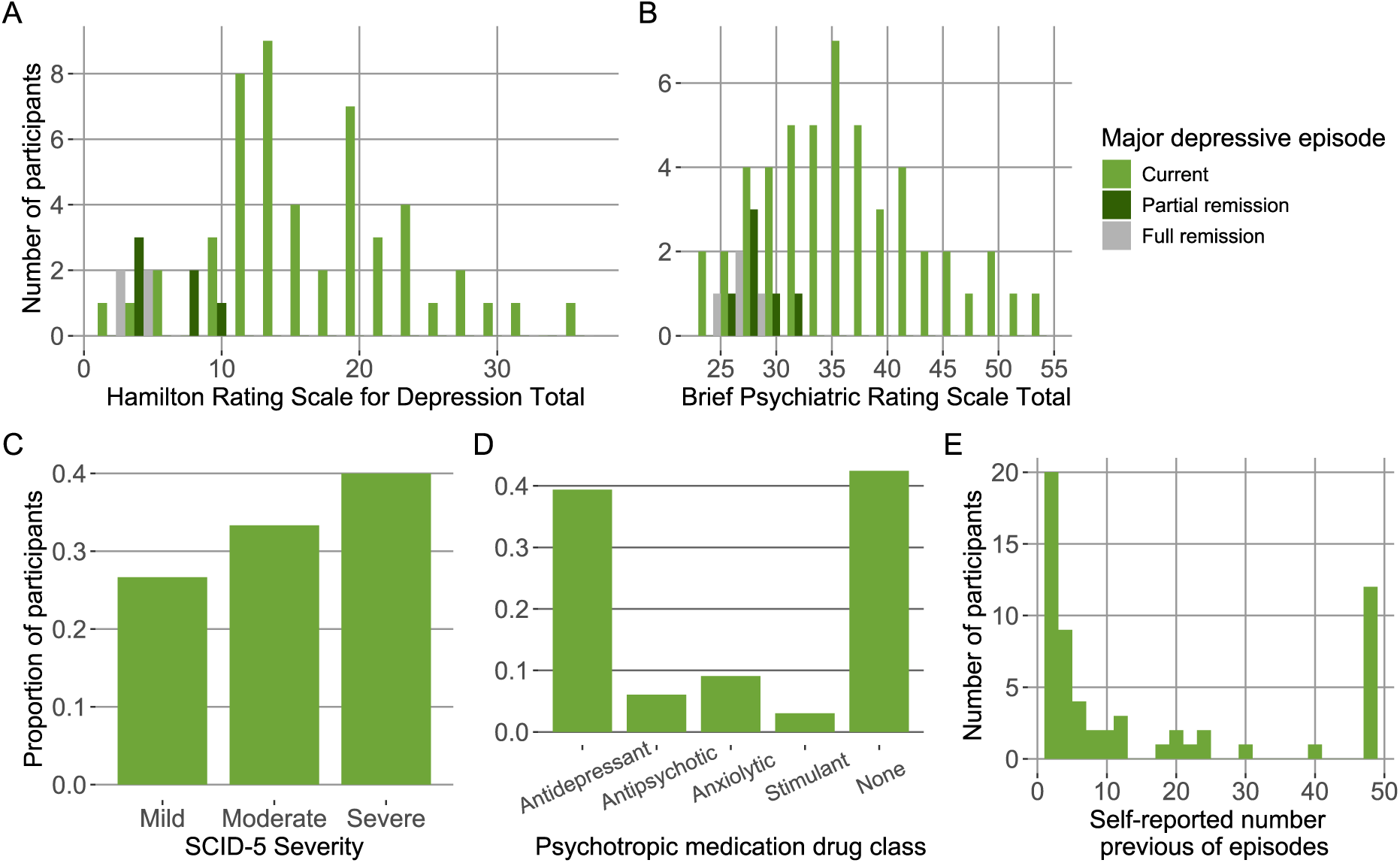
MDD sample characteristics. Histograms, A & B, y-axis: number of MDD participants, x-axis: A: Hamilton Depression Rating Scale total, B: Brief Psychiatric Rating Scale total, fill indicates major depressive episode status (light green indicates current, dark green indicates partial remission, and grey indicates full remission). C, D, & E, y-axis: proportion of MDD participants, C: Structured Clinical Interview for DSM-5 severity rating, D: psychotropic medication drug class, E: self-reported previous number of major depressive episodes.

**Figure S.I. 5:**
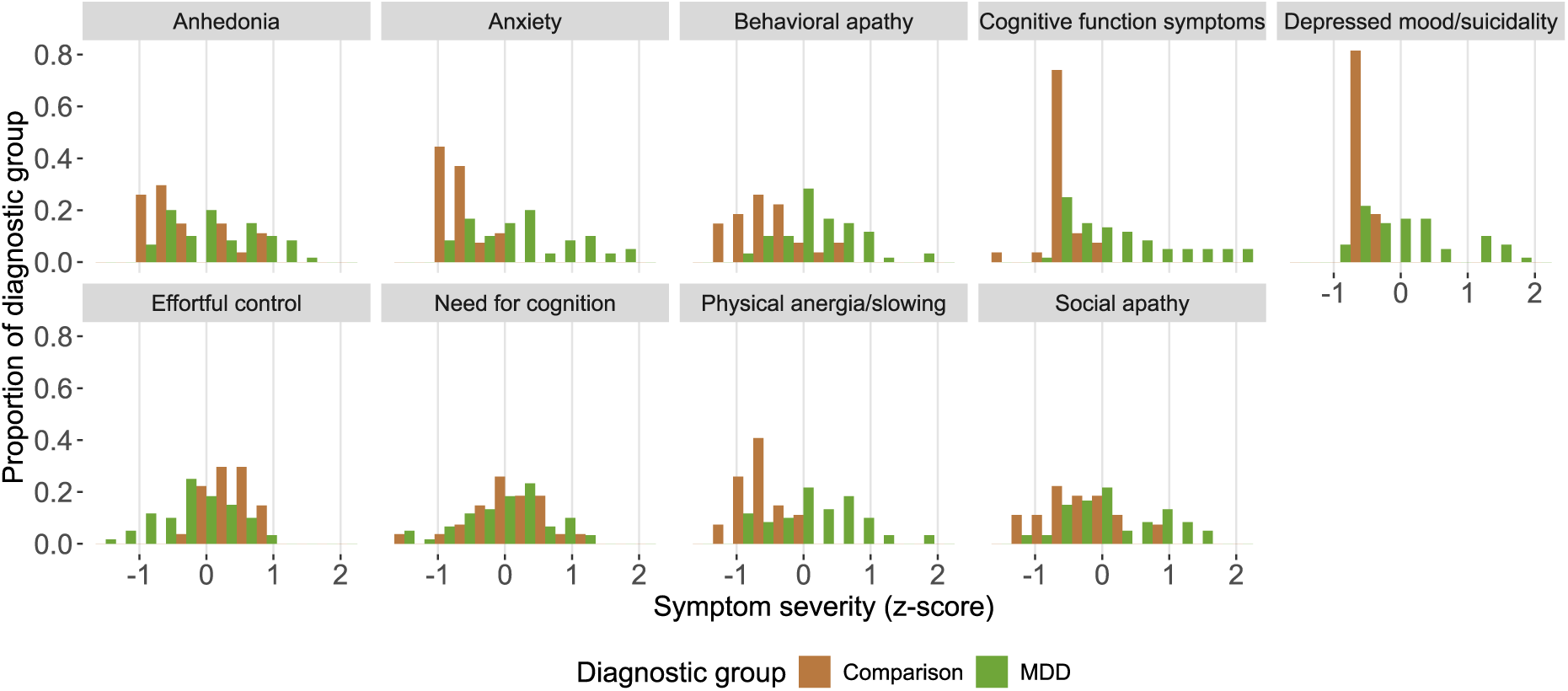
Symptom severity by domain and diagnostic group. Histogram, bar color indicates diagnostic group.

**Figure S.I. 6:**
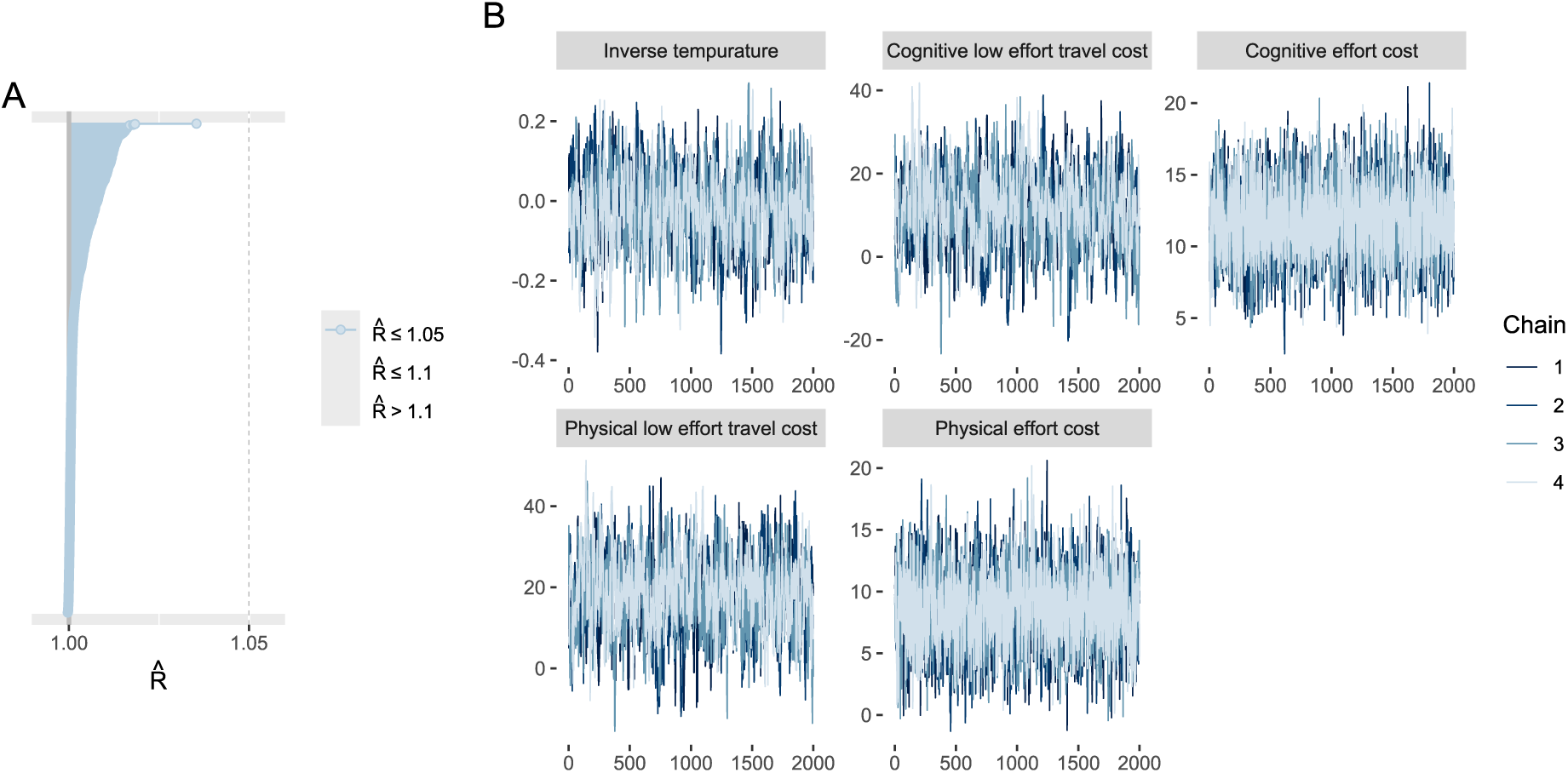
MVT model diagnostics. A: R^^^ convergence diagnostic plot for all parameters, all of which are below the 1.05 cutoff (vertical dotted line) indicating model convergence. B: MCMC trace plots, color indicates chain number, overlapping traces suggests model convergence.

**Figure S.I. 7:**
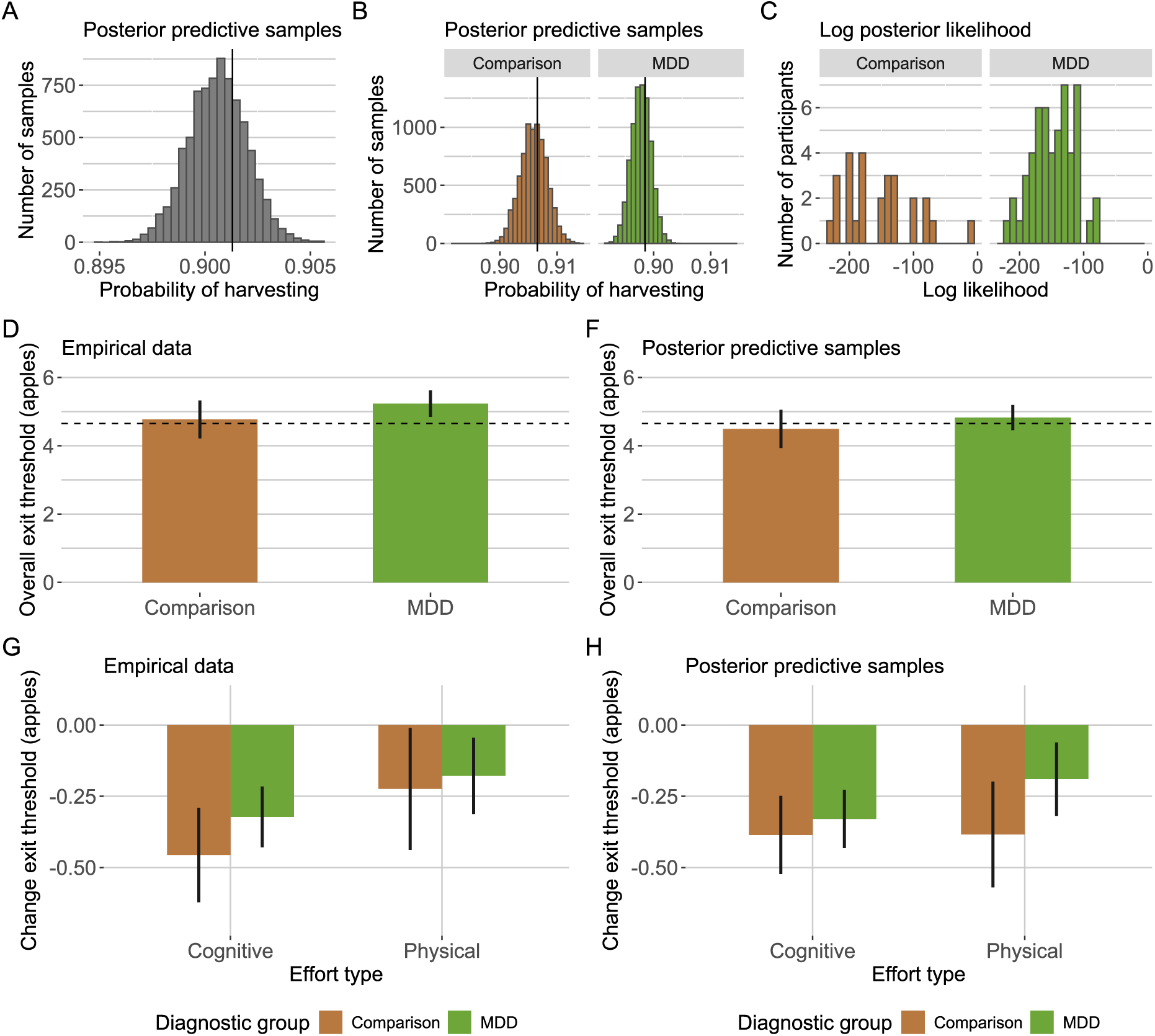
Posterior predictive check results by diagnostic group. A: x-axis, probability of choosing to harvest (1) or exit (0) across all trials, y-axis, number of MCMC samples, empirical observation indicated by vertical black line. B: x-axis, probability of choosing to harvest (1) or exit (0) across all trials for comparison participants (left) and MDD participants (right), y-axis, number of MCMC samples, empirical observations indicated by vertical black lines. C: x-axis, log posterior likelihood per participant for comparison (left) and MDD participants (right), y-axis number of participants. D: empirical data, x-axis, diagnostic group, y-axis, overall exit threshold (from low effort orchards), bars indicate group means, error bars indicate standard error of the mean, horizontal dotted line indicates best threshold from simulation. D: posterior predictive data, for each MCMC sample we computed the overall exit threshold per participant, and aggregated across samples to get the mean value per participant. Resulting plot shows group means and SEM matching C. E. empirical data, x-axis, effort type, y-axis, change exit threshold (high - low effort orchards), bars indicate group means, error bars indicate standard error of the mean, fill indicates diagnostic group. F. posterior predictive data, for each MCMC sample we computed the change in exit threshold per participant, and aggregated across samples to get the mean value per participant. Resulting plot shows group means and SEM matching E.

**Figure S.I. 8:**
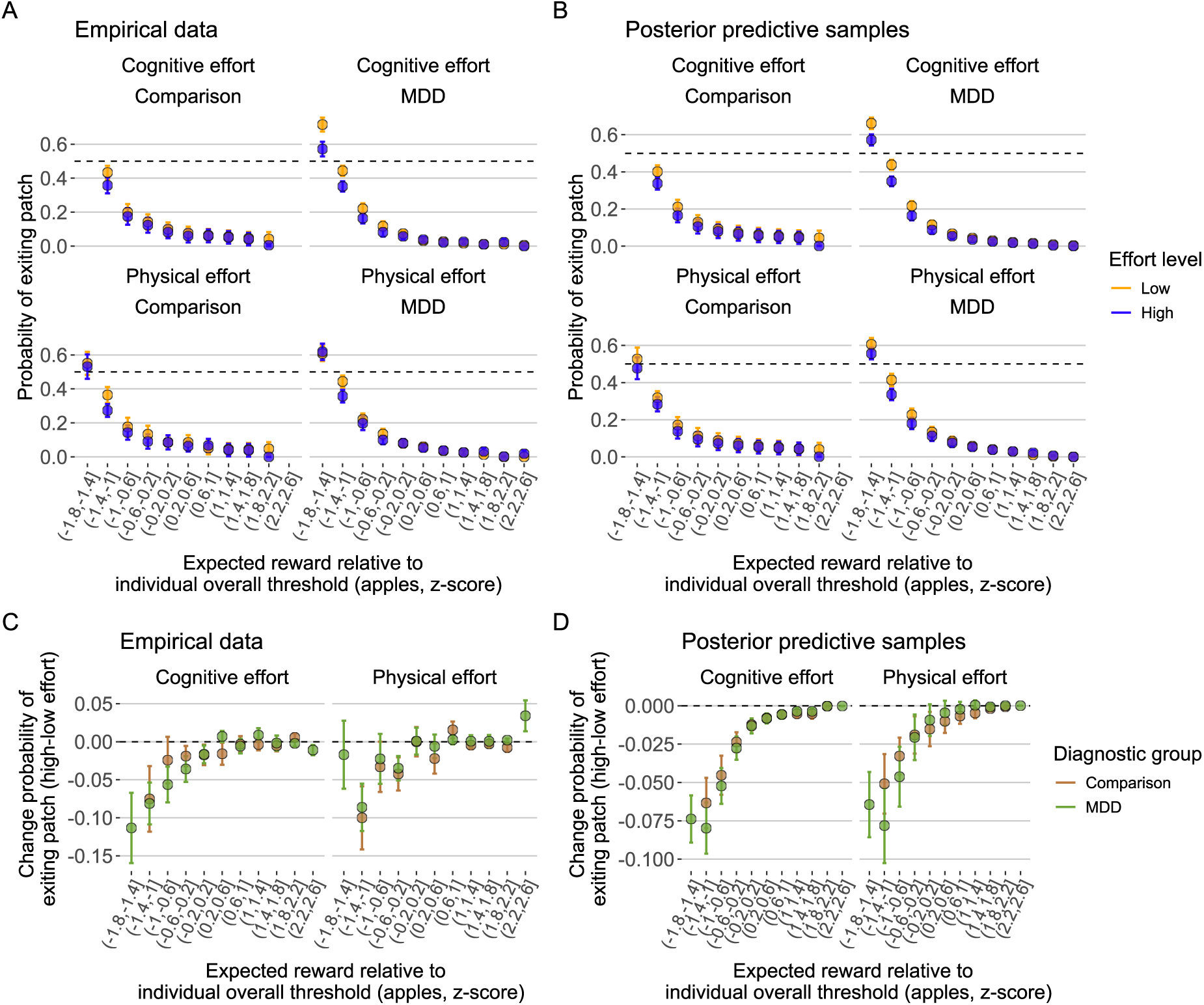
Probability of exiting by expected reward level, posterior predictive check results. A: empirical data, x-axis indicates relative expected reward, within each participant computed their overall exit threshold from the average of the low effort orchards using mixed-effects linear regression, then computed the difference between expected reward on each trial and the overall exit threshold (i.e., ’relative expected reward’), then z-scored this difference within participant and then binned the z-scores by 0.4 z-score units. y-axis indicates the probability of exiting the patch in each bin, points indicate group-level mean and error bars indicate standard error of the mean, points with fewer than 20 observations not displayed. First row indicates cognitive effort, and second row indicates physical effort, first column indicates comparison group, and second column indicates MDD group. B: posterior predictive data, same as panel A, except for each MCMC sample we computed the probability of exiting in each of the relative expected reward bins, and aggregated across samples to get the mean value per participant. Plot shows group-level means and SEM matching panel A. C. empirical data, x-axis, relative expected reward binned, y-axis, change in the probability of exiting patch (high - low effort orchards), bars indicate group-level means, error bars indicate standard error of the mean, fill indicates diagnostic group, brown color indicates comparison group, green color indicates MDD group, points with fewer than 25 observations not displayed. D. posterior predictive data, same as panel A, except for each MCMC sample we computed the change probability of exiting patch per participant, and aggregated across samples to get the mean value per participant. Resulting plot shows group means and SEM matching panel C.

**Figure S.I. 9:**
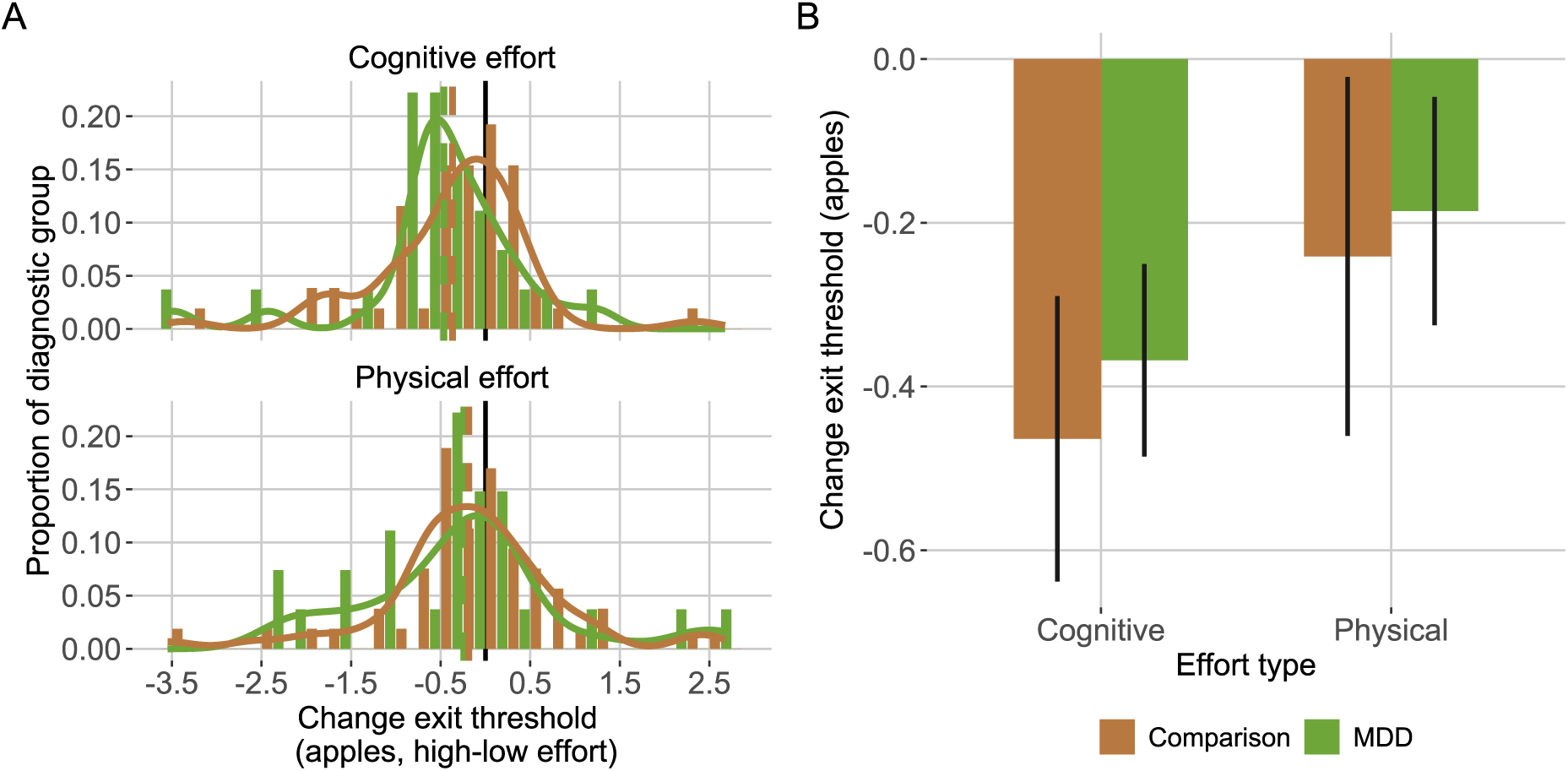
Model-agnostic change in exit threshold by diagnostic group. A: individual differences, error bars indicate 95% HDI, x-axis indicates cognitive effort condition, y-axis indicates physical effort condition. B: diagnostic group differences, x-axis indicates effort type and diagnostic group, y-axis indicates change in the exit threshold (high - low effort, apples).

**Figure S.I. 10:**
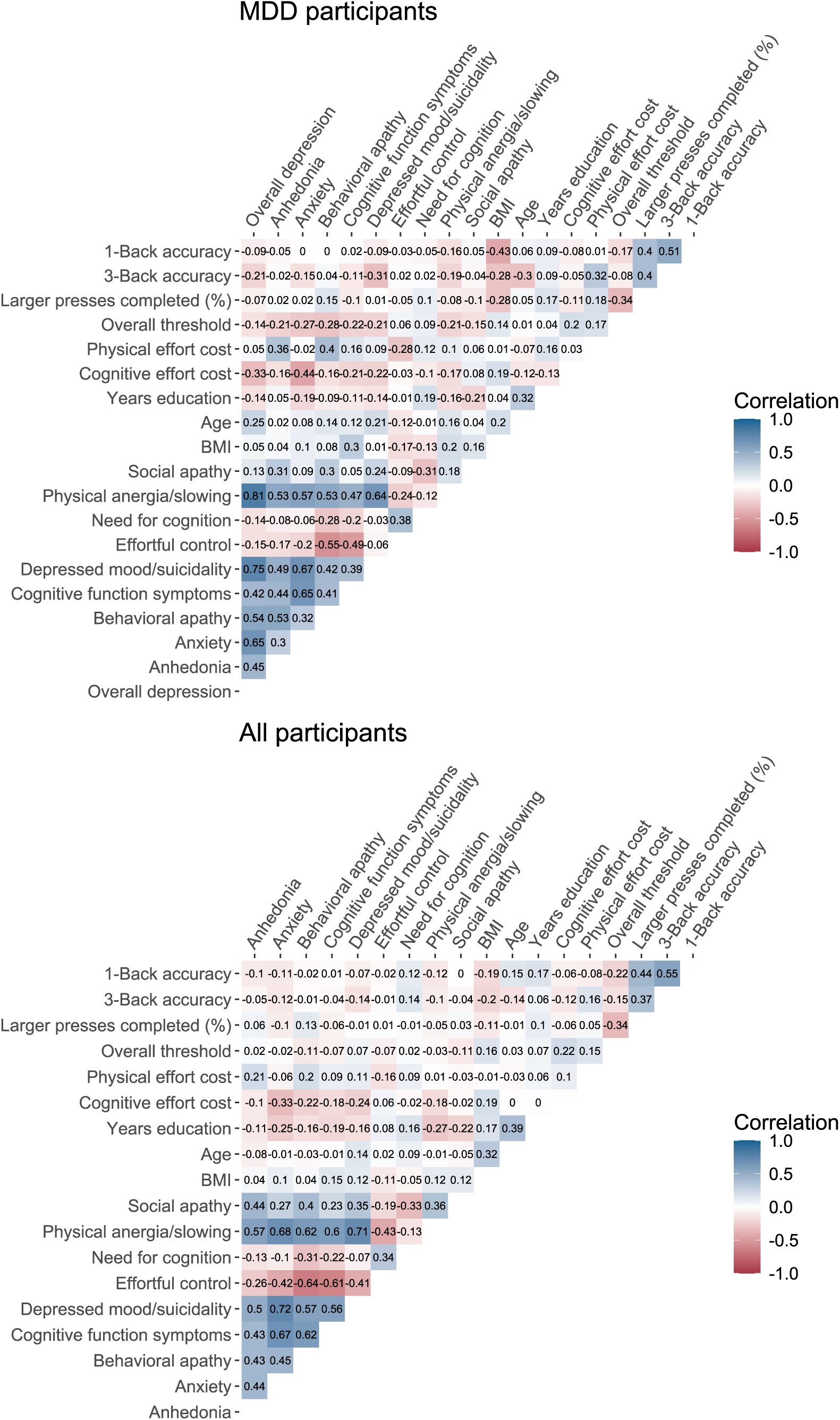
Task behavior symptom heatmap. Top MDD group only, bottom all participants (spearman correlation matrix).

**Figure S.I. 11:**
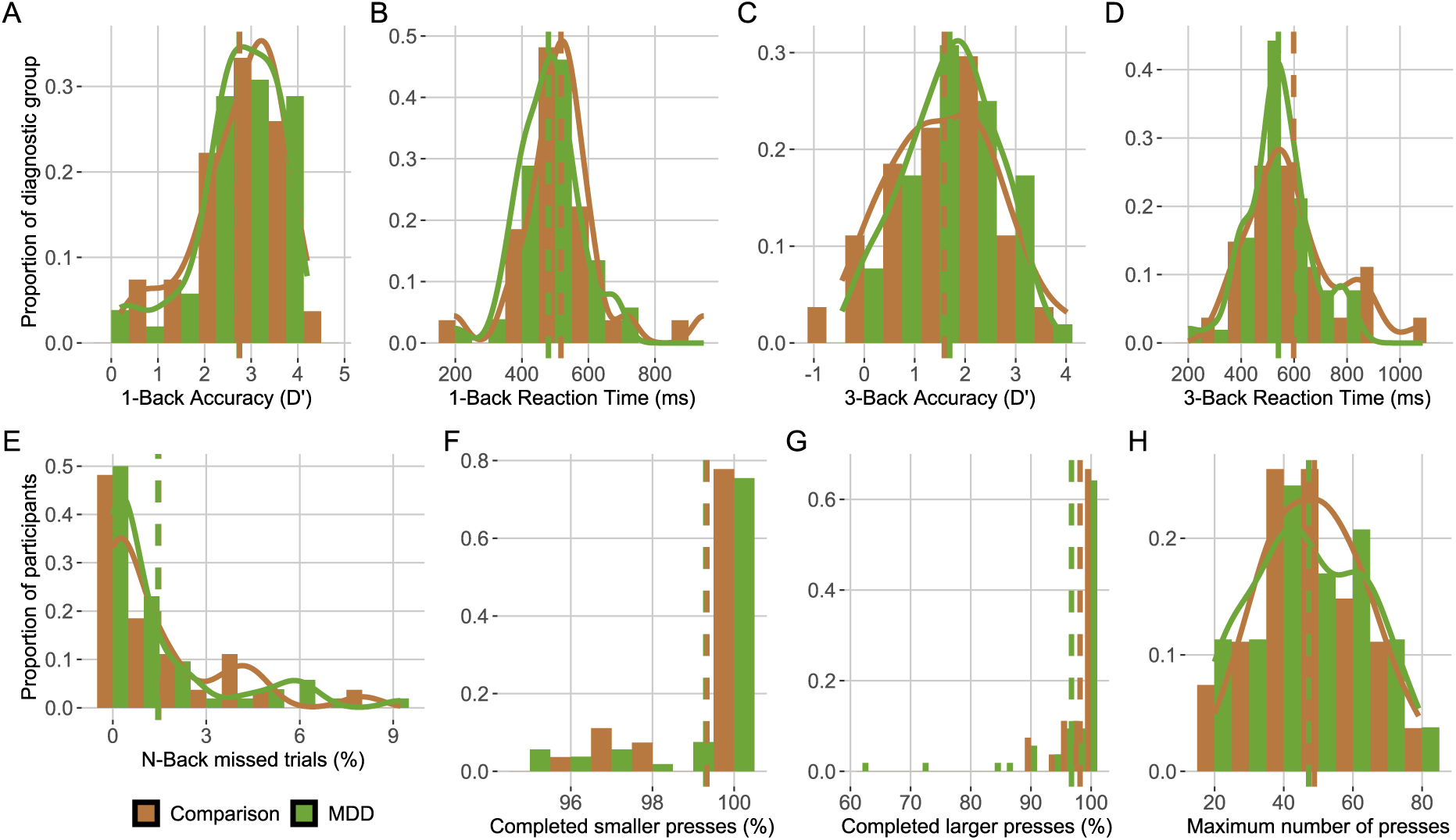
Diagnostic group difference in travel task performance. A: 1-Back accuracy (D’), B: 1-Back Reaction Time (ms), C: 3-Back accuracy (D’), D: 3-Back Reaction Time (ms), E: percent of N-Back trials missed responding before the RT deadline, F: Physical low effort performance (percent completed smaller number of presses), G: Physical high effort performance (percent completed larger number of presses), H: Maximum number of presses determined in a calibration phase. There were no diagnostic group differences except that the MDD group responded faster on average on the cognitive (N-Back) task (A & C), and completed fewer presses in the high effort physical condition (G).

**Figure S.I. 12:**
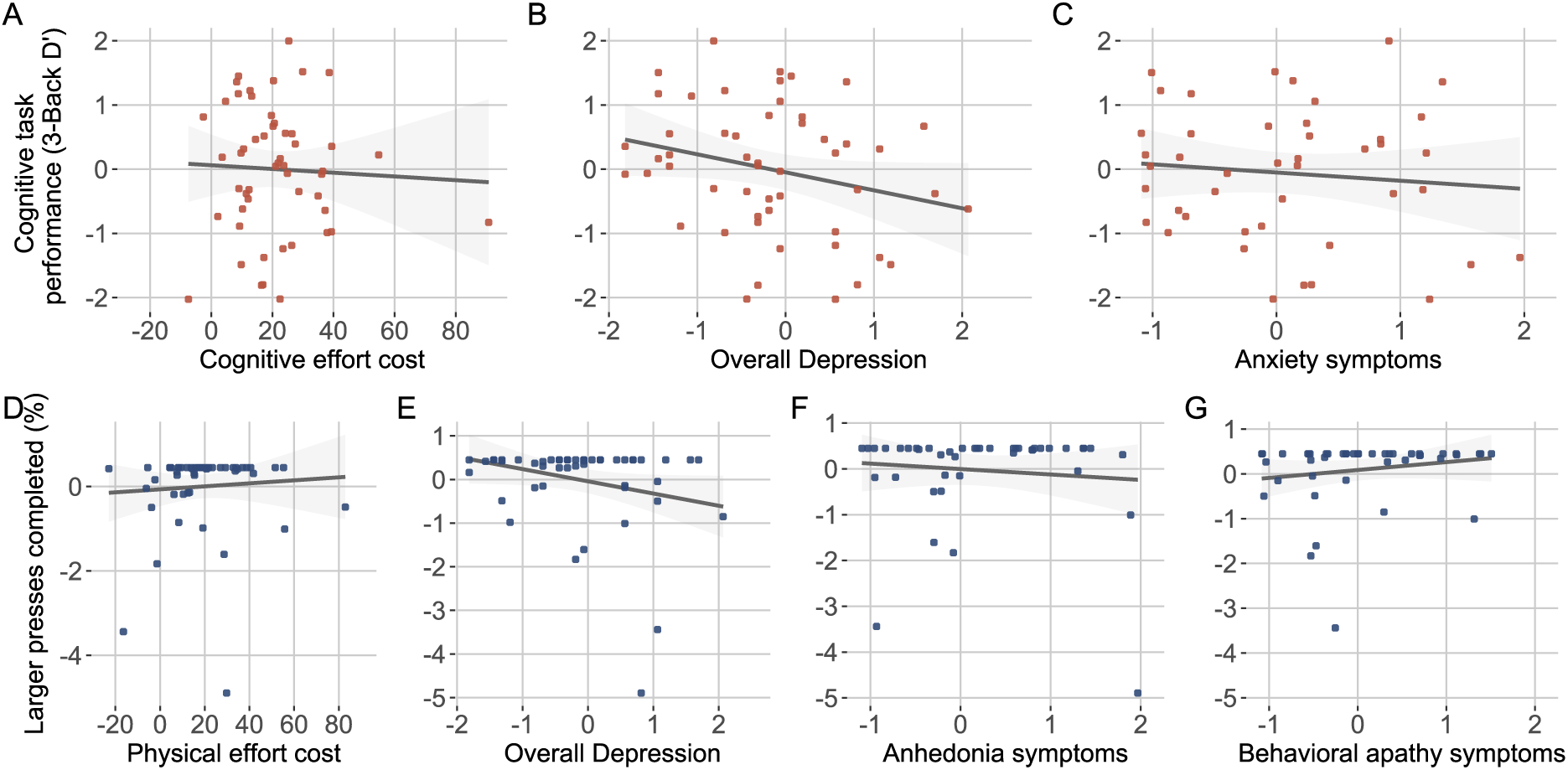
Travel task performance relationship to effort costs, overall depression, anhedonia, and anxiety. Only MDD group. Cognitive travel task performance (3-Back Accuracy (D’), y-axis) not reliably related to, A: cognitive effort cost (*p>*0.470), B: overall depression (HAMD, z-score, *p>*0.16), nor C: anxiety symptoms (x-axis, z-score, *p>*0.21). Physical travel task performance (percent of larger number of presses completed, y-axis) not reliably related to, D: physical effort cost (*p>*0.828), E: overall depression (*p>*0.073), nor F: anhedonia symptoms (x-axis, *p>*0.318), nor G: behavioral apathy symptoms (*p>*0.240).

**Table S.I. 1:** Clinical interview and self-report battery. Clinical interview and self-reports were completed in session 1 in the order listed in this table. Scales asking about similar timescales were grouped together.

| Scale name | N items | Abbreviation | Time scale |
| --- | --- | --- | --- |
| Hamilton Depression Rating Scale | 21 | HAM-D | Past week |
| Brief Psychiatric Rating Scale | 24 | BPRS | Last 2 weeks |
| MGH Cognitive and Physical Functioning Questionnaire | 7 | MGH-CPFQ | Last month |
| Patient Health Questionnaire-9 | 9 | PHQ-9 | Last 2 weeks |
| Generalized Anxiety Disorder-7 | 7 | GAD7 | Last 2 weeks |
| Apathy Motivation Index | 14 | AMI | Last 2 weeks |
| Snaith-Hamilton Pleasure Scale | 13 | SHAPS | Last few days |
| Adult Temperament Questionnaire-Effortful Control | 19 | ATQ-EC | Trait |
| Need for Cognition | 18 | NFC | Trait |
| **Total Self-report** | 87 |  |  |

**Table S.I. 2:** Confirmatory symptom factors alpha and diagnostic group differences. Column 1: symptom domain, column 2: Cronbach’s alpha (95% confidence interval), column 3: number of items, column 4-8: MDD mean, Comparison mean, and t-statistic, degrees of freedom, p-value. Emotional apathy and appetite symptoms had low Cronbach’s alpha scores and were not included in further analysis.

| Symptom domain | alpha(95% CI) | N | MDD | Comp. | *t* | df | *p* |
| --- | --- | --- | --- | --- | --- | --- | --- |
| Anhedonia | 0.89(0.84-0.92) | 15 | 0.132 | -0.294 | 3.07 | 55.46 | 0.003 |
| Anxiety | 0.90(0.86-0.93) | 10 | 0.197 | -0.626 | 7.386 | 83.523 | <0.001 |
| Appetite symptoms | 0.52(0.20-0.69) | 3 | 0.091 | -0.609 | 5.501 | 76.74 | <0.001 |
| Behavioral apathy | 0.72(0.53-0.82) | 8 | 0.186 | -0.554 | 6.124 | 58.79 | <0.001 |
| Emotional apathy | 0.54(0.30-0.68) | 5 | -0.049 | 0.137 | -1.22 | 44.08 | 0.230 |
| Social apathy | 0.71(0.55-0.81) | 5 | 0.149 | -0.331 | 3.303 | 58.024 | 0.002 |
| Cognitive function symp. | 0.84(0.75-0.89) | 5 | 0.257 | -0.572 | 6.880 | 80.293 | <0.001 |
| Depressed mood | 0.86(0.81-0.90) | 8 | 0.106 | -0.626 | 7.508 | 66.346 | <0.001 |
| Effortful control (trait) | 0.84(0.76-0.88) | 17 | -0.177 | 0.393 | -6.29 | 80.5 | <0.001 |
| Need for cognition (trait) | 0.90(0.85-0.93) | 17 | -0.033 | 0.074 | -0.794 | 58.151 | 0.430 |
| Physical anergia/slowing | 0.76(0.64-0.84) | 7 | 0.157 | -0.611 | 7.616 | 84.89 | <0.001 |

**Table S.I. 3:**
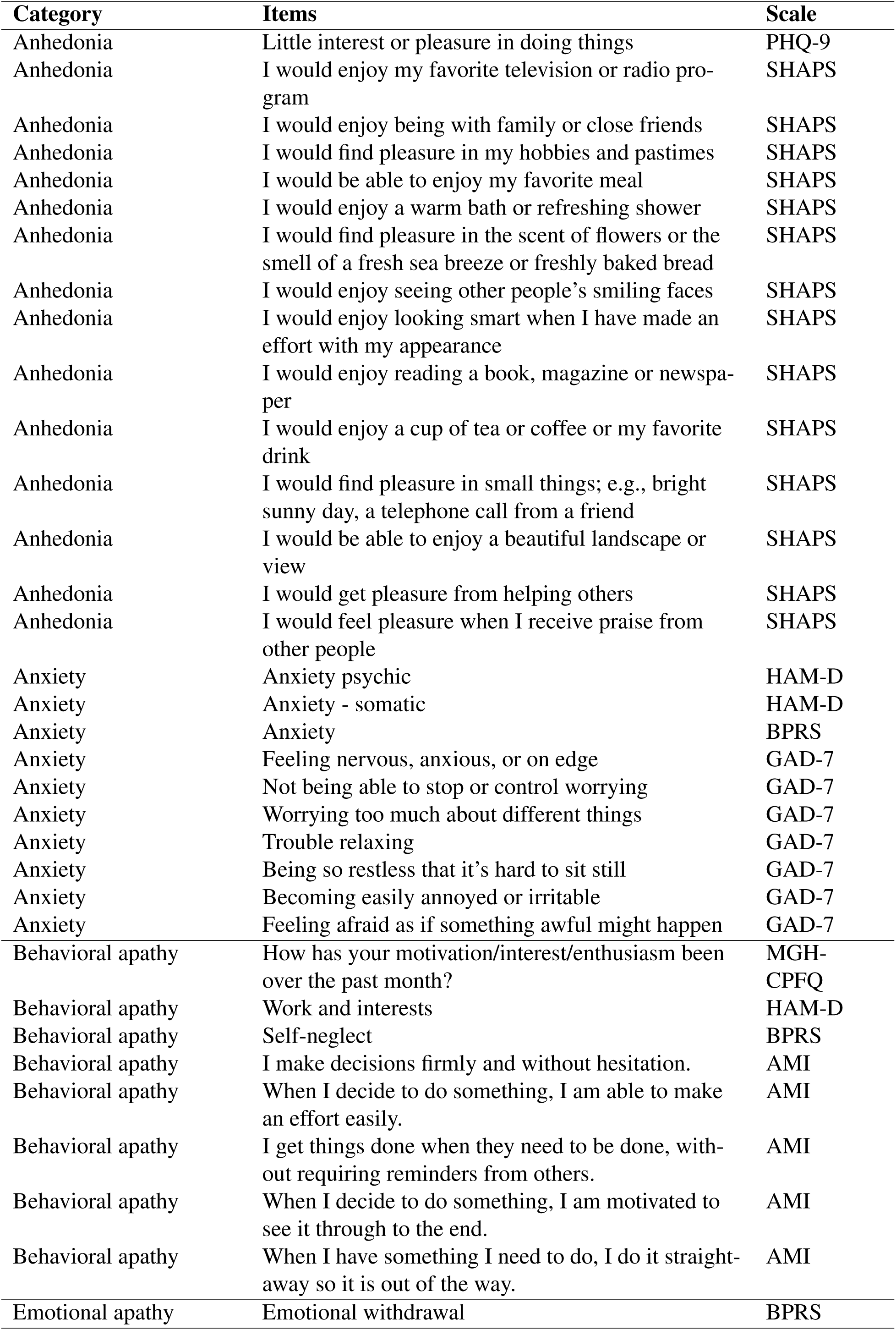
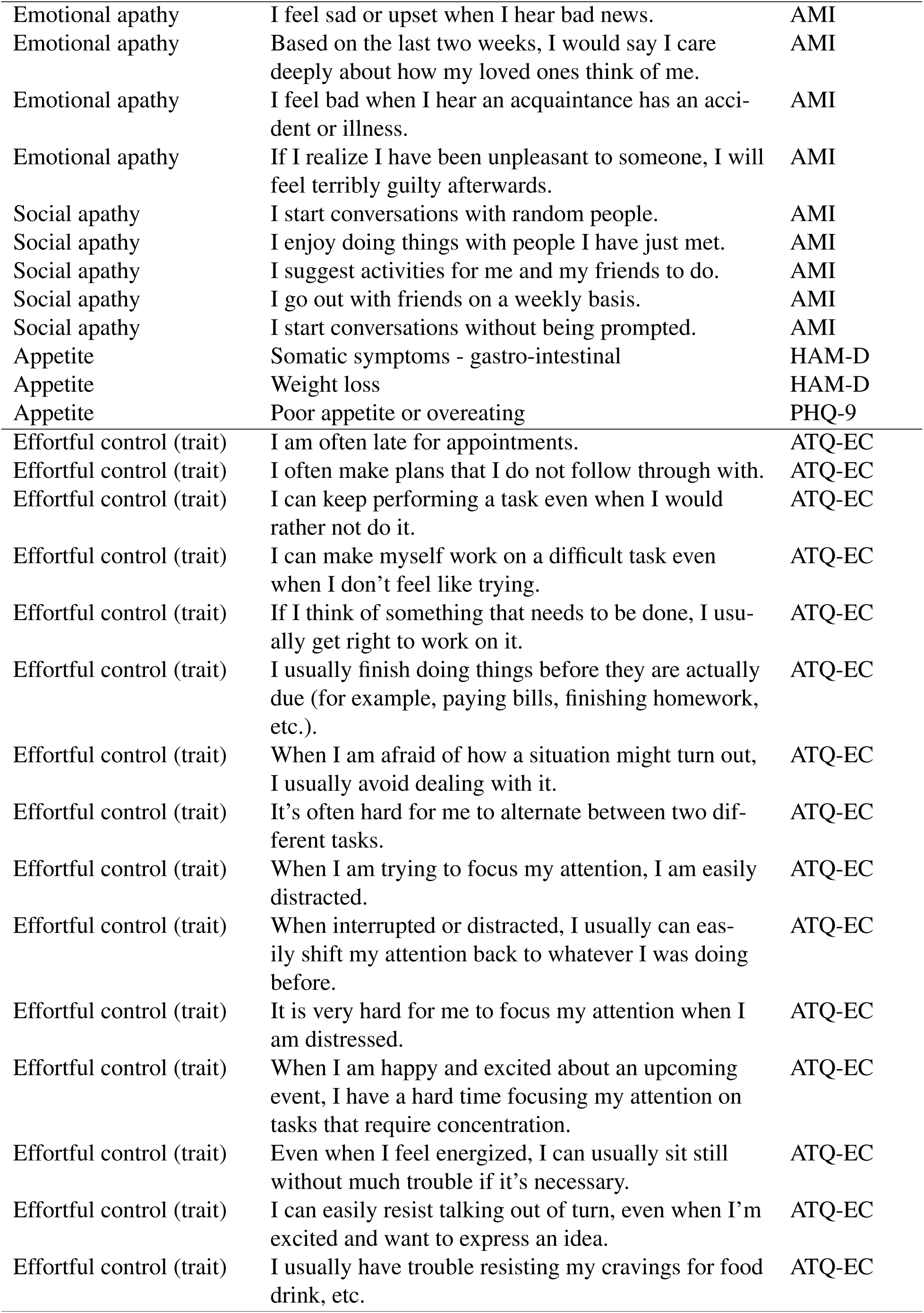
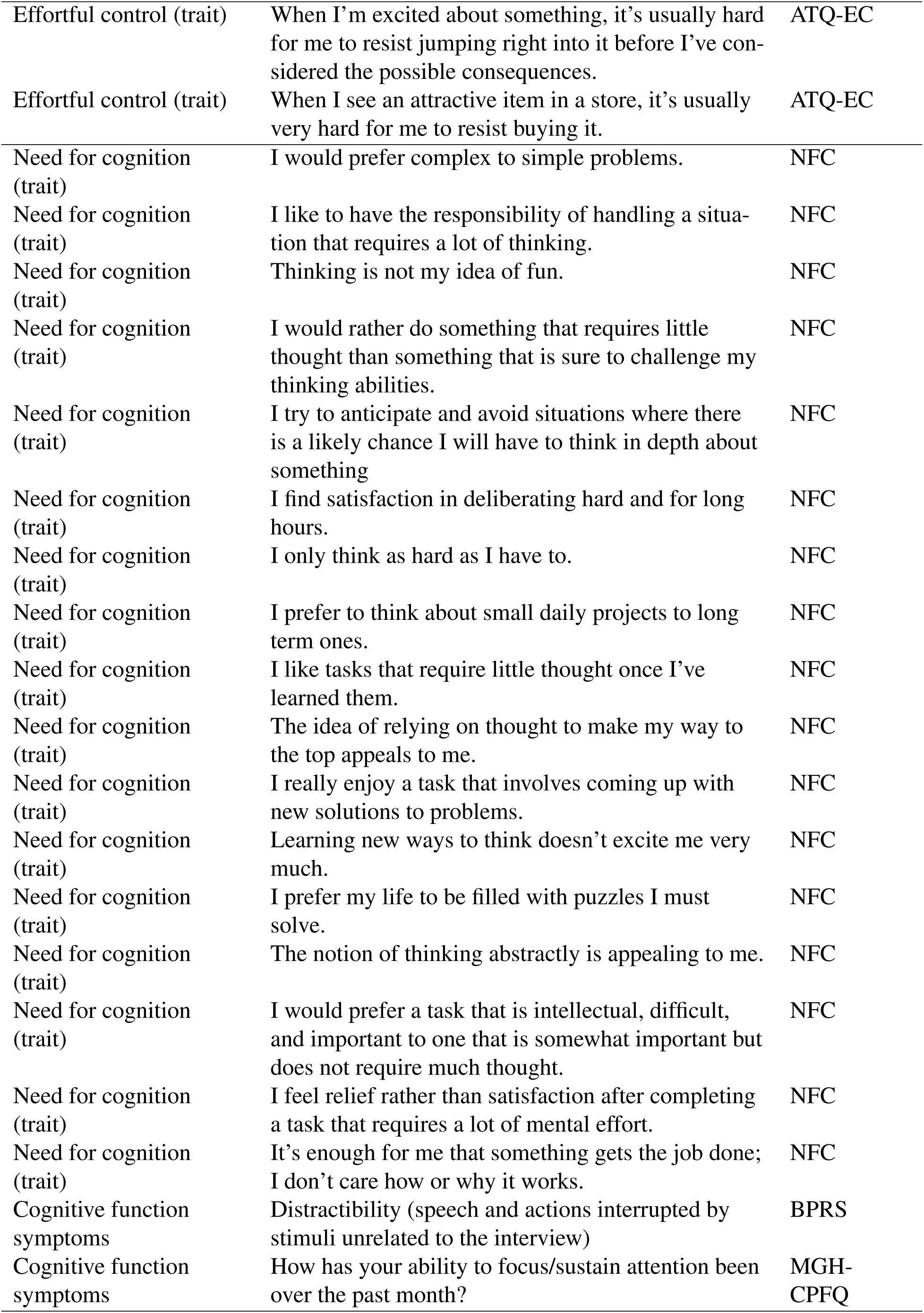
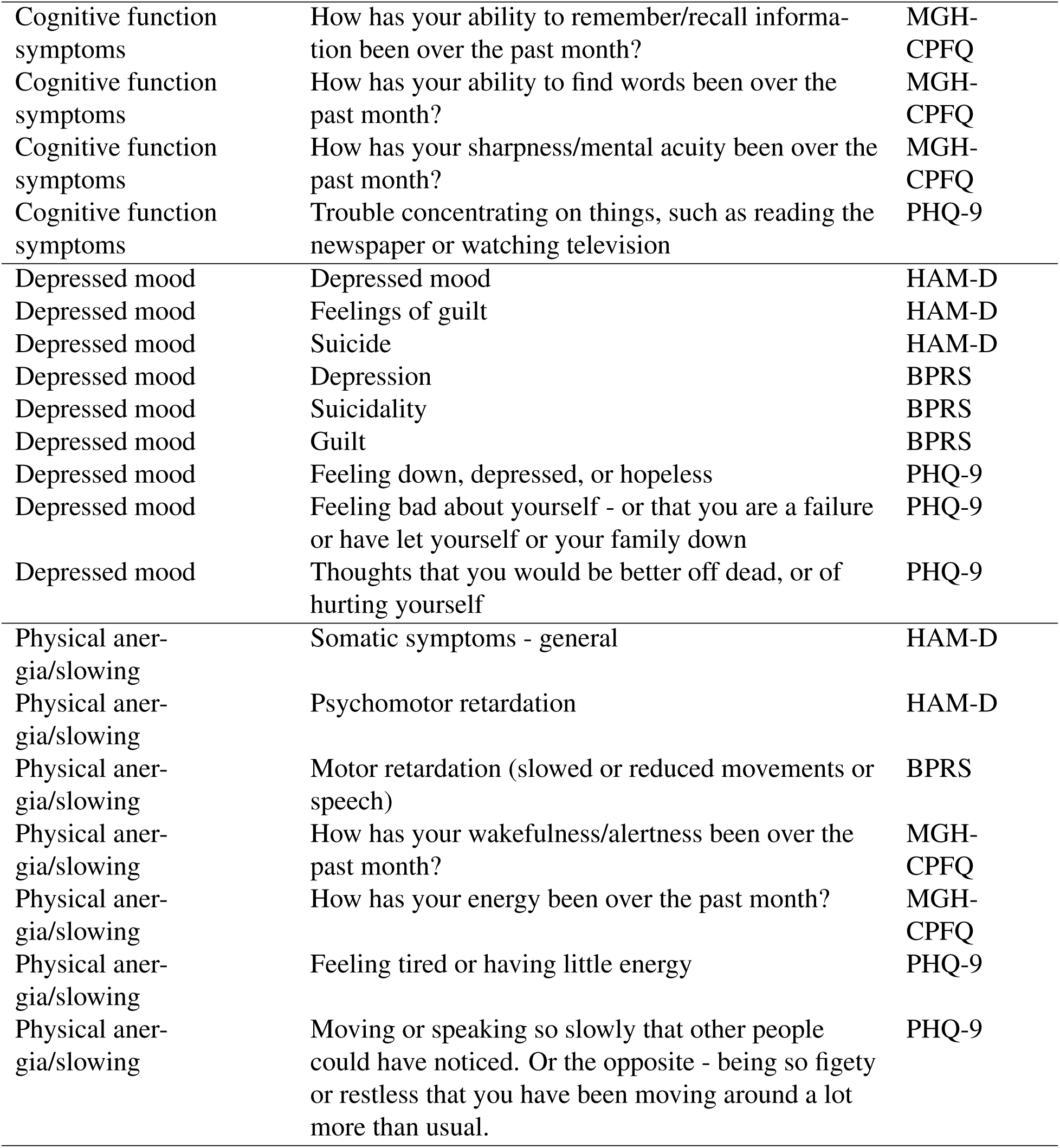
Items for confirmatory symptom factors. Column 1: symptom domain, column 2: items, column 3: measurement scale (abbreviations in Table S.I. 1).

Major Depression Severity and Effort in Foraging Supplementary Information
| Category | Items | Scale |
| --- | --- | --- |
| Anhedonia | Little interest or pleasure in doing things | PHQ-9 |
| Anhedonia | I would enjoy my favorite television or radio program | SHAPS |
| Anhedonia | I would enjoy being with family or close friends | SHAPS |
| Anhedonia | I would find pleasure in my hobbies and pastimes | SHAPS |
| Anhedonia | I would be able to enjoy my favorite meal | SHAPS |
| Anhedonia | I would enjoy a warm bath or refreshing shower | SHAPS |
| Anhedonia | I would find pleasure in the scent of flowers or the smell of a fresh sea breeze or freshly baked bread | SHAPS |
| Anhedonia | I would enjoy seeing other people's smiling faces | SHAPS |
| Anhedonia | I would enjoy looking smart when I have made an effort with my appearance | SHAPS |
| Anhedonia | I would enjoy reading a book, magazine or newspaper | SHAPS |
| Anhedonia | I would enjoy a cup of tea or coffee or my favorite drink | SHAPS |
| Anhedonia | I would find pleasure in small things; e.g., bright sunny day, a telephone call from a friend | SHAPS |
| Anhedonia | I would be able to enjoy a beautiful landscape or view | SHAPS |
| Anhedonia | I would get pleasure from helping others | SHAPS |
| Anhedonia | I would feel pleasure when I receive praise from other people | SHAPS |
| Anxiety | Anxiety psychic | HAM-D |
| Anxiety | Anxiety - somatic | HAM-D |
| Anxiety | Anxiety | BPRS |
| Anxiety | Feeling nervous, anxious, or on edge | GAD-7 |
| Anxiety | Not being able to stop or control worrying | GAD-7 |
| Anxiety | Worrying too much about different things | GAD-7 |
| Anxiety | Trouble relaxing | GAD-7 |
| Anxiety | Being so restless that it's hard to sit still | GAD-7 |
| Anxiety | Becoming easily annoyed or irritable | GAD-7 |
| Anxiety | Feeling afraid as if something awful might happen | GAD-7 |
| Behavioral apathy | How has your motivation/interest/enthusiasm been over the past month? | MGH-CPFQ |
| Behavioral apathy | Work and interests | HAM-D |
| Behavioral apathy | Self-neglect | BPRS |
| Behavioral apathy | I make decisions firmly and without hesitation. | AMI |
| Behavioral apathy | When I decide to do something, I am able to make an effort easily. | AMI |
| Behavioral apathy | I get things done when they need to be done, without requiring reminders from others. | AMI |
| Behavioral apathy | When I decide to do something, I am motivated to see it through to the end. | AMI |
| Behavioral apathy | When I have something I need to do, I do it straight-away so it is out of the way. | AMI |
| Emotional apathy | Emotional withdrawal | BPRS |

| Category | Items | Scale |
| --- | --- | --- |
| Emotional apathy | I feel sad or upset when I hear bad news. | AMI |
| Emotional apathy | Based on the last two weeks, I would say I care deeply about how my loved ones think of me. | AMI |
| Emotional apathy | I feel bad when I hear an acquaintance has an accident or illness. | AMI |
| Emotional apathy | If I realize I have been unpleasant to someone, I will feel terribly guilty afterwards. | AMI |
| Social apathy | I start conversations with random people. | AMI |
| Social apathy | I enjoy doing things with people I have just met. | AMI |
| Social apathy | I suggest activities for me and my friends to do. | AMI |
| Social apathy | I go out with friends on a weekly basis. | AMI |
| Social apathy | I start conversations without being prompted. | AMI |
| Appetite | Somatic symptoms - gastro-intestinal | HAM-D |
| Appetite | Weight loss | HAM-D |
| Appetite | Poor appetite or overeating | PHQ-9 |
| Effortful control (trait) | I am often late for appointments. | ATQ-EC |
| Effortful control (trait) | I often make plans that I do not follow through with. | ATQ-EC |
| Effortful control (trait) | I can keep performing a task even when I would rather not do it. | ATQ-EC |
| Effortful control (trait) | I can make myself work on a difficult task even when I don't feel like trying. | ATQ-EC |
| Effortful control (trait) | If I think of something that needs to be done, I usually get right to work on it. | ATQ-EC |
| Effortful control (trait) | I usually finish doing things before they are actually due (for example, paying bills, finishing homework, etc.). | ATQ-EC |
| Effortful control (trait) | When I am afraid of how a situation might turn out, I usually avoid dealing with it. | ATQ-EC |
| Effortful control (trait) | It's often hard for me to alternate between two different tasks. | ATQ-EC |
| Effortful control (trait) | When I am trying to focus my attention, I am easily distracted. | ATQ-EC |
| Effortful control (trait) | When interrupted or distracted, I usually can easily shift my attention back to whatever I was doing before. | ATQ-EC |
| Effortful control (trait) | It is very hard for me to focus my attention when I am distressed. | ATQ-EC |
| Effortful control (trait) | When I am happy and excited about an upcoming event, I have a hard time focusing my attention on tasks that require concentration. | ATQ-EC |
| Effortful control (trait) | Even when I feel energized, I can usually sit still without much trouble if it's necessary. | ATQ-EC |
| Effortful control (trait) | I can easily resist talking out of turn, even when I'm excited and want to express an idea. | ATQ-EC |
| Effortful control (trait) | I usually have trouble resisting my cravings for food drink, etc. | ATQ-EC |
| Effortful control (trait) | When I'm excited about something, it's usually hard for me to resist jumping right into it before I've considered the possible consequences. | ATQ-EC |
| Effortful control (trait) | When I see an attractive item in a store, it's usually very hard for me to resist buying it. | ATQ-EC |
| Need for cognition (trait) | I would prefer complex to simple problems. | NFC |
| Need for cognition (trait) | I like to have the responsibility of handling a situation that requires a lot of thinking. | NFC |
| Need for cognition (trait) | Thinking is not my idea of fun. | NFC |
| Need for cognition (trait) | I would rather do something that requires little thought than something that is sure to challenge my thinking abilities. | NFC |
| Need for cognition (trait) | I try to anticipate and avoid situations where there is a likely chance I will have to think in depth about something | NFC |
| Need for cognition (trait) | I find satisfaction in deliberating hard and for long hours. | NFC |
| Need for cognition (trait) | I only think as hard as I have to. | NFC |
| Need for cognition (trait) | I prefer to think about small daily projects to long term ones. | NFC |
| Need for cognition (trait) | I like tasks that require little thought once I've learned them. | NFC |
| Need for cognition (trait) | The idea of relying on thought to make my way to the top appeals to me. | NFC |
| Need for cognition (trait) | I really enjoy a task that involves coming up with new solutions to problems. | NFC |
| Need for cognition (trait) | Learning new ways to think doesn't excite me very much. | NFC |
| Need for cognition (trait) | I prefer my life to be filled with puzzles I must solve. | NFC |
| Need for cognition (trait) | The notion of thinking abstractly is appealing to me. | NFC |
| Need for cognition (trait) | I would prefer a task that is intellectual, difficult, and important to one that is somewhat important but does not require much thought. | NFC |
| Need for cognition (trait) | I feel relief rather than satisfaction after completing a task that requires a lot of mental effort. | NFC |
| Need for cognition (trait) | It's enough for me that something gets the job done; I don't care how or why it works. | NFC |
| Cognitive function symptoms | Distractibility (speech and actions interrupted by stimuli unrelated to the interview) | BPRS |
| Cognitive function symptoms | How has your ability to focus/sustain attention been over the past month? | MGH-CPFQ |
| Cognitive function symptoms | How has your ability to remember/recall information been over the past month? | MGH-CPFQ |
| Cognitive function symptoms | How has your ability to find words been over the past month? | MGH-CPFQ |
| Cognitive function symptoms | How has your sharpness/mental acuity been over the past month? | MGH-CPFQ |
| Cognitive function symptoms | Trouble concentrating on things, such as reading the newspaper or watching television | PHQ-9 |
| Depressed mood | Depressed mood | HAM-D |
| Depressed mood | Feelings of guilt | HAM-D |
| Depressed mood | Suicide | HAM-D |
| Depressed mood | Depression | BPRS |
| Depressed mood | Suicidality | BPRS |
| Depressed mood | Guilt | BPRS |
| Depressed mood | Feeling down, depressed, or hopeless | PHQ-9 |
| Depressed mood | Feeling bad about yourself - or that you are a failure or have let yourself or your family down | PHQ-9 |
| Depressed mood | Thoughts that you would be better off dead, or of hurting yourself | PHQ-9 |
| Physical anergia/slowing | Somatic symptoms - general | HAM-D |
| Physical anergia/slowing | Psychomotor retardation | HAM-D |
| Physical anergia/slowing | Motor retardation (slowed or reduced movements or speech) | BPRS |
| Physical anergia/slowing | How has your wakefulness/alertness been over the past month? | MGH-CPFQ |
| Physical anergia/slowing | How has your energy been over the past month? | MGH-CPFQ |
| Physical anergia/slowing | Feeling tired or having little energy | PHQ-9 |
| Physical anergia/slowing | Moving or speaking so slowly that other people could have noticed. Or the opposite - being so fidgety or restless that you have been moving around a lot more than usual. | PHQ-9 |

**Table S.I. 4:** Demographic factors by diagnostic group. Column 1: demographic factor and response options. Continuous measures shown first with means (columns 2-3) and unpaired t-test statistics (columns 4-6, * indicates *p<*0.05). Years of education (self) was significantly different between groups, but not correlated with effort costs. Diagnostic groups were matched on all other variables, and this was also true within the subset of participants that were included in the task behavioral analyses (statistical tests denoted by ’behavioral analyses’).

|  | **MDD** |  | **Comparison** |  |
| --- | --- | --- | --- | --- |
|  | **Mean** | **SD(range)** | **Mean** | **SD(range)** |
| **Age** ($t=-0.083$, $df=57.2$, $p=0.934$) | | | | |
|  | 26.92 | 11.1(18-61) | 27.11 | 9.64(19-59) |
| **Age, behavioral analyses** ($t=-0.47$, $df=53.94$, $p=0.643$) | | | | |
| **Childhood income** ($t=-2.35$, $df=61.73$, $p=0.022$) | | | | |
|  | 5.07 | 1.98(1-8) | 6.00 | 1.52(3-9) |
| **Childhood income, behavioral analyses** ($t=-2.12$, $df=62.94$, $p=0.038$) | | | | |
| **Years education** |  |  |  |  |
| **Mother** ($t=0.086$, $df=61.8$, $p=0.931$) | | | | |
|  | 15.13 | 3.41(5-20) | 15.07 | 2.73(11-20) |
| **Mother, behavioral analyses** ($t=0.46$, $df=62.41$, $p=0.645$) | | | | |
| **Father** ($t=-1.17$, $df=61.93$, $p=0.245$) | | | | |
|  | 14.8 | 4.08(0-20) | 15.8 | 3.26(12-20) |
| **Father, behavioral analyses** ($t=-0.75$, $df=59.07$, $p=0.456$) | | | | |
| **Self** ($t=-2.51$, $df=47.7$, $p<0.016^*$) | | | | |
|  | 14.7 | 2.15(9-21) | 16.0 | 2.28(13-20) |
| **Self, behavioral analyses** ($t=-2.40$, $df=49.17$, $p<0.020$) | | | | |
| **Education-self correlation with effort costs** |  |  |  |  |
|  | **Correlation** | **t** | **df** | **p** |
| Cognitive effort cost, MDD group | -0.03 | -0.24 | 50 | 0.809 |
| Cognitive effort cost, all participants | 0.09 | 0.80 | 77 | 0.427 |
| Physical effort cost, MDD group | 0.15 | 1.07 | 51 | 0.290 |
| Physical effort cost, all participants) | 0.06 | 0.53 | 78 | 0.599 |
|  | **MDD** |  | **Comparison** |  |
|  | **N** | **%** | **N** | **%** |
| **Gender** ($\chi^2=2.39$, $df=2$, $p=0.303$) | | | | |
| **Gender, behavioral analyses** ($\chi^2=3.75$, $df=2$, $p=0.154$) | | | | |
| Female | 38 | 63.3% | 15 | 55.6% |
| Male | 19 | 31.7% | 12 | 44.4% |
| Non-binary | 3 | 5% | 0 | 0% |
| **Race** ($\chi^2=1.66$, $df=4$, $p=0.800$) | | | | |
| **Race, behavioral analyses** ($\chi^2=1.63$, $df=4$, $p=0.803$) | | | | |
| American Indian/Alaska Native | 0 | 0% | 0 | 0% |
| Asian | 13 | 21.7% | 8 | 29.6% |
| Black or African American | 8 | 13.3% | 3 | 11.1% |
| Native Hawaiian/Pacific Islander | 0 | 0% | 0 | 0% |
| White | 27 | 45% | 13 | 48.1% |
| Other or prefer not to say | 6 | 10% | 2 | 7.41% |
| More than one race | 6 | 10% | 1 | 3.7% |
| **Ethnicity** ($\chi^2=1.41$, $df=2$, $p=0.495$) | | | | |
| **Ethnicity, behavioral analyses** ($\chi^2=1.71$, $df=2$, $p=0.424$) | | | | |
| Latino or Hispanic | 8 | 13.3% | 4 | 14.8% |
|  | **N** | **%** | **N** | **%** |
| Not Latino or Hispanic | 49 | 81.7% | 23 | 85.2% |
| Other or prefer not to say | 3 | 5% | 0 | 0% |
| **Total household income** ($\chi^2=9.44$, $df=7$, $p=0.222$) | | | | |
| **Total household income, behavioral analyses** ($\chi^2=9.02$, $df=7$, $p=0.251$) | | | | |
| Less than \$25,000 | 12 | 20% | 1 | 3.7% |
| \$25,000 to \$34,999 | 7 | 11.7% | 3 | 11.1% |
| \$35,000 to \$49,999 | 8 | 13.3% | 2 | 7.41% |
| \$50,000 to \$74,999 | 9 | 15% | 8 | 29.6% |
| \$75,000 to \$99,999 | 5 | 8.33% | 5 | 18.5% |
| \$100,000 to \$149,999 | 6 | 10% | 3 | 11.1% |
| \$150,000 to \$199,999 | 0 | 0% | 1 | 3.7% |
| \$200,000 or more | 4 | 6.67% | 2 | 7.41% |
| Prefer not to answer | 9 | 15% | 2 | 7.41% |
| **Occupational status** ($\chi^2=9.64$, $df=7$, $p=0.209$) | | | | |
| **Occupational status, behavioral analyses** ($\chi^2=10.39$, $df=7$, $p=0.167$) | | | | |
| Working full-time | 9 | 15% | 11 | 40.7% |
| Working part-time | 7 | 11.7% | 2 | 7.41% |
| Student full-time | 9 | 15% | 3 | 11.1% |
| Student part-time | 3 | 5% | 0 | 0% |
| Working & student | 21 | 35% | 9 | 33.3% |
| Homemaker | 2 | 3.33% | 0 | 0% |
| Retired | 0 | 0% | 0 | 0% |
| Volunteer worker | 0 | 0% | 0 | 0% |
| Seeking employment | 6 | 10% | 2 | 7.41% |
| Leave of absence | 0 | 0% | 0 | 0% |
| Disabled (other free response) | 3 | 5% | 0 | 0% |
| **Relationship status** ($\chi^2=2.94$, $df=3$, $p=0.400$) | | | | |
| **Relationship status, behavioral analyses** ($\chi^2=4.12$, $df=3$, $p=0.248$) | | | | |
| Single | 48 | 80% | 21 | 77.8% |
| Married | 2 | 3.33% | 3 | 11.1% |
| Divorced or separated | 2 | 3.33% | 0 | 0% |
| Widowed | 0 | 0% | 0 | 0% |
| Other | 8 | 13.3% | 3 | 11.1% |
| **Alcohol frequency** ($\chi^2=2.20$, $df=4$, $p=0.700$) | | | | |
| **Alcohol frequency, behavioral analyses** ($\chi^2=2.27$, $df=4$, $p=0.686$) | | | | |
| Never | 15 | 25% | 8 | 29.6% |
| Monthly or less | 14 | 23.3% | 7 | 25.9% |
| 2-4 times a month | 20 | 33.3% | 5 | 18.5% |
| 2-3 times a week | 9 | 15% | 6 | 22.2% |
| 4 or more times a week | 2 | 3.33% | 1 | 3.7% |
| **Alcohol amount** ($\chi^2=2.46$, $df=4$, $p=0.651$) | | | | |
| **Alcohol amount, behavioral analyses** ($\chi^2=2.19$, $df=4$, $p=0.702$) | | | | |
| 0 (N/A) | 15 | 25% | 8 | 29.6% |
| 1 or 2 | 20 | 33.3% | 10 | 37.0% |
| 3 or 4 | 16 | 26.7% | 5 | 18.5% |
| 5 or 6 | 6 | 10.0% | 4 | 14.8% |

Table S.I. 4: Demographic factors by diagnostic group.
|  | ***MDD*** |  | ***Comparison*** |  |
| --- | --- | --- | --- | --- |
|  | **N** | **%** | **N** | **%** |
| 7, 8, or 9 | 3 | 5.0% | 0 | 0% |
| 10 or more | 0 | 0% | 0 | 0% |
| ***Caffeine amount*** ($\chi^2=2.59$, $df=4$, $p=0.623$) | | | | |
| ***Caffeine amount, behavioral analyses*** ($\chi^2=2.04$, $df=3$, $p=0.565$) | | | | |
| None | 15 | 25% | 7 | 25.9% |
| 1 cup | 29 | 48.3% | 11 | 40.7% |
| 2-3 cups | 13 | 21.7% | 9 | 33.3% |
| 4-5 cups | 2 | 3.33% | 0 | 0% |
| 6 or more cups | 1 | 1.67% | 0 | 0% |
| ***Tobacco*** ($\chi^2=2.71$, $df=1$, $p=0.099$) | | | | |
| ***Tobacco, behavioral analyses*** ($\chi^2=2.31$, $df=1$, $p=0.129$) | | | | |
| Uses tobacco | 12 | 20% | 1 | 3.7% |
| Does not use tobacco | 48 | 80% | 26 | 96.3% |

**Table S.I. 5:** Group-level parameter posterior distribution values. Column 1: parameter, column 2: mean of the group-level posterior distribution, column 3: lower bound of 95% credible interval, column 4: upper bound of credible interval, column 5: Bayesian p-values.

| Parameter | Mean | Lower bound | Upper bound | *p* |
| --- | --- | --- | --- | --- |
| Inverse Temperature (log) | -0.02 | -0.20 | 0.15 | 0.408 |
| Cognitive Low Effort Travel Cost | 10.97 | -6.69 | 27.19 | 0.098 |
| Cognitive Effort Cost | 11.71 | 7.10 | 16.47 | <0.001 |
| Physical Low Effort Travel Cost | 18.18 | 0.00 | 35.06 | 0.025 |
| Physical Effort Cost | 8.59 | 3.03 | 14.21 | 0.001 |
| Cognitive vs. Physical Effort Cost Correlation | 0.05 | -0.24 | 0.34 | >0.365 |

**Table S.I. 6:** Symptoms effort cost regressions, controlling for psychotropic medication use, MDD participants. (A, MDD) Predict cognitive effort cost by overall depression severity and each symptom domain, controlling for cognitive task performance (3-Back D’), medication use, age, and years of education. (B, MDD) Predict physical effort cost by overall depression severity, and each symptom domain, controlling for physical task performance (% larger number of presses completed), BMI, medication use, age and years of education. (C) All participants, predict overall exit threshold (log apples, from low effort conditions) by diagnostic group (MDD-comparison) and psychotropic medication use. MDD participants, predict over-all exit threshold (log apples, from low effort conditions) by symptom severity, medication use, age, and years of education (* indicates *p<*0.05, FDR correction within symptom models). All variables were scaled as input to the regressions.

| Symptom | Estimate | SE | *t* | *p* | *p<sub>adjusted</sub>* |
| --- | --- | --- | --- | --- | --- |
| **A. Cognitive effort cost, MDD, controlling for medication use** |  |  |  |  |  |
| Diagnostic group difference | -0.02 | 0.29 | -0.08 | 0.935 |  |
| Overall depression | -0.39 | 0.16 | -2.35 | 0.023* |  |
| Anhedonia | -0.20 | 0.15 | -1.32 | 0.192 | 0.227 |
| Anxiety | -0.49 | 0.14 | -3.62 | 0.001 | 0.005* |
| Behavioral apathy | -0.20 | 0.15 | -1.32 | 0.195 | 0.227 |
| Social apathy | 0.01 | 0.15 | 0.06 | 0.953 | 0.953 |
| Cognitive function symptoms | -0.30 | 0.14 | -2.11 | 0.041 | 0.144 |
| Depressed mood/suicidality | -0.26 | 0.17 | -1.55 | 0.129 | 0.226 |
| Physical anergia/slowness | -0.24 | 0.15 | -1.59 | 0.118 | 0.226 |
| **B. Physical effort cost, MDD, controlling for medication use** |  |  |  |  |  |
| Diagnostic group difference | 0.12 | 0.29 | 0.40 | 0.692 |  |
| Overall depression | 0.04 | 0.17 | 0.22 | 0.828 |  |
| Anhedonia | 0.42 | 0.14 | 2.90 | 0.006 | 0.035* |
| Anxiety | -0.04 | 0.16 | -0.25 | 0.804 | 0.804 |
| Behavioral apathy | 0.40 | 0.15 | 2.68 | 0.010 | 0.035* |
| Social apathy | 0.14 | 0.15 | 0.93 | 0.360 | 0.491 |
| Cognitive function symptoms | 0.27 | 0.16 | 1.65 | 0.106 | 0.247 |
| Depressed mood/suicidality | 0.14 | 0.17 | 0.81 | 0.421 | 0.491 |
| Physical anergia/slowness | 0.16 | 0.17 | 0.98 | 0.331 | 0.491 |
| **C. Overall exit threshold, MDD, controlling for medication use** |  |  |  |  |  |
| Diagnostic group difference | 0.57 | 0.28 | 2.04 | 0.045* |  |
| Psychotropic medication use | -0.568 | 0.274 | -2.074 | 0.0415* |  |
| Overall depression | -0.38 | 0.15 | -2.47 | 0.017* |  |
| Anhedonia | -0.29 | 0.14 | -2.15 | 0.037 | 0.052 |
| Anxiety | -0.33 | 0.14 | -2.36 | 0.022 | 0.052 |
| Behavioral apathy | -0.31 | 0.14 | -2.22 | 0.031 | 0.052 |
| Social apathy | -0.11 | 0.14 | -0.82 | 0.414 | 0.41 |
| Cognitive function symptoms | -0.22 | 0.14 | -1.54 | 0.129 | 0.151 |
| Depressed mood/suicidality | -0.40 | 0.15 | -2.73 | 0.009 | 0.047* |
| Physical anergia/slowness | -0.35 | 0.14 | -2.57 | 0.013 | 0.047* |

**Table S.I. 7:** Diagnostic group difference MVT model. Group effect parameter for model that included all MDD participants. Current depressed group effect parameter for model that excluded participants in remission.

| **Diagnostic group effect parameter** | Mean | Lower | Upper | *pd* |
| --- | --- | --- | --- | --- |
| Cognitive Effort Cost | -1.03 | -8.05 | 6.08 | 0.39 |
| Physical Effort Cost | -1.03 | -8.05 | 6.08 | 0.39 |
| Cognitive Low Effort Cost | 1.81 | -7.30 | 10.72 | 0.34 |
| Physical Low Effort Cost | -3.06 | -12.02 | 5.90 | 0.26 |
| Inverse temperature (log) | 0.10 | -0.24 | 0.46 | 0.29 |
| **Current depressed group effect parameter** | Mean | Lower | Upper | *p* |
| Cognitive Effort Cost | -1.39 | -8.49 | 5.76 | 0.35 |
| Physical Effort Cost | -1.73 | -9.10 | 5.60 | 0.32 |
| Cognitive Low Effort Cost | 2.52 | -6.53 | 11.77 | 0.29 |
| Physical Low Effort Cost | -3.73 | -13.01 | 5.40 | 0.21 |
| Inverse temperature (log) | 0.09 | -0.29 | 0.49 | 0.32 |

**Table S.I. 8:** Self-reported overall depression regression results. Column 1: predicted variable, self-reported overall depression (PHQ-9) was used as a predictor variable in place of clinician-rated depression (HAMD). Results correspond to patterns identified using clinician rated depression. All variables were scaled as input to the regressions.

| Predicted variable | Estimate | SE | *t* | *p* |
| --- | --- | --- | --- | --- |
| Cognitive effort cost | -0.32 | 0.15 | -2.13 | 0.038* |
| Physical effort cost | 0.08 | 0.15 | 0.52 | 0.606 |
| 1-Back D' | -0.16 | 0.17 | -0.92 | 0.361 |
| 3-Back D' | -0.12 | 0.18 | -0.66 | 0.512 |
| Smaller number of presses completed (%) | 0.01 | 0.15 | 0.06 | 0.951 |
| Larger number of presses completed (%) | -0.14 | 0.16 | -0.91 | 0.369 |
| Overall exit threshold | -0.44 | 0.14 | -3.15 | 0.003* |

**Table S.I. 9:** Symptoms effort cost regressions, current MDD only, and all participants. (A, current MDD, C, all participants) Predict cognitive effort cost by overall depression severity and each symptom domain, controlling for cognitive task performance (3-Back D’), age, and years of education. (B, current MDD, D, all participants) Predict physical effort cost by overall depression severity, and each symptom domain, controlling for physical task performance (% larger number of presses completed), BMI, age and years of education (* indicates *p<*0.05, FDR correction within symptom models). All variables were scaled as input to the regressions.

| Symptom | Estimate | SE | *t* | *p* | *p<sub>adjusted</sub>* |
| --- | --- | --- | --- | --- | --- |
| **A. Cognitive effort cost, current MDD participants** |  |  |  |  |  |
| Overall depression | -0.29 | 0.21 | -1.38 | 0.176 |  |
| Anhedonia | -0.13 | 0.17 | -0.78 | 0.443 | 0.517 |
| Anxiety | -0.50 | 0.15 | -3.32 | 0.002 | 0.014* |
| Behavioral apathy | -0.21 | 0.16 | -1.32 | 0.195 | 0.456 |
| Social apathy | -0.07 | 0.17 | -0.44 | 0.664 | 0.664 |
| Cognitive function symptoms | -0.33 | 0.17 | -1.99 | 0.054 | 0.189 |
| Depressed mood/suicidality | -0.20 | 0.18 | -1.11 | 0.273 | 0.477 |
| Physical anergia/slowness | -0.17 | 0.18 | -0.93 | 0.357 | 0.500 |
| **B. Physical effort cost, current MDD participants** |  |  |  |  |  |
| Overall depression | 0.30 | 0.19 | 1.55 | 0.131 |  |
| Anhedonia | 0.54 | 0.13 | 4.06 | <0.001 | <0.001* |
| Anxiety | 0.06 | 0.16 | 0.35 | 0.729 | 0.851 |
| Behavioral apathy | 0.41 | 0.14 | 3.01 | 0.005 | 0.018* |
| Social apathy | 0.02 | 0.15 | 0.14 | 0.888 | 0.888 |
| Cognitive function symptoms | 0.45 | 0.16 | 2.81 | 0.008 | 0.019* |
| Depressed mood/suicidality | 0.23 | 0.16 | 1.43 | 0.163 | 0.228 |
| Physical anergia/slowness | 0.36 | 0.17 | 2.16 | 0.038 | 0.066 |
| **C. Cognitive effort cost, all participants** |  |  |  |  |  |
| Anhedonia | -0.13 | 0.12 | -1.11 | 0.268 | 0.313 |
| Anxiety | -0.40 | 0.12 | -3.44 | 0.001 | 0.007* |
| Behavioral apathy | -0.21 | 0.12 | -1.74 | 0.086 | 0.120 |
| Social apathy | -0.02 | 0.12 | -0.15 | 0.880 | 0.880 |
| Cognitive function symptoms | -0.24 | 0.12 | -2.02 | 0.047 | 0.120 |
| Depressed mood/suicidality | -0.23 | 0.13 | -1.75 | 0.085 | 0.120 |
| Physical anergia/slowness | -0.21 | 0.12 | -1.75 | 0.085 | 0.120 |
| **D. Physical effort cost, all participants** |  |  |  |  |  |
| Anhedonia | 0.27 | 0.12 | 2.30 | 0.024 | 0.168 |
| Anxiety | -0.04 | 0.13 | -0.32 | 0.753 | 0.835 |
| Behavioral apathy | 0.21 | 0.12 | 1.71 | 0.091 | 0.318 |
| Social apathy | 0.03 | 0.12 | 0.21 | 0.835 | 0.835 |
| Cognitive function symptoms | 0.14 | 0.12 | 1.19 | 0.239 | 0.558 |
| Depressed mood/suicidality | 0.10 | 0.14 | 0.74 | 0.459 | 0.803 |
| Physical anergia/slowness | 0.06 | 0.13 | 0.45 | 0.654 | 0.835 |

**Table S.I. 10:**
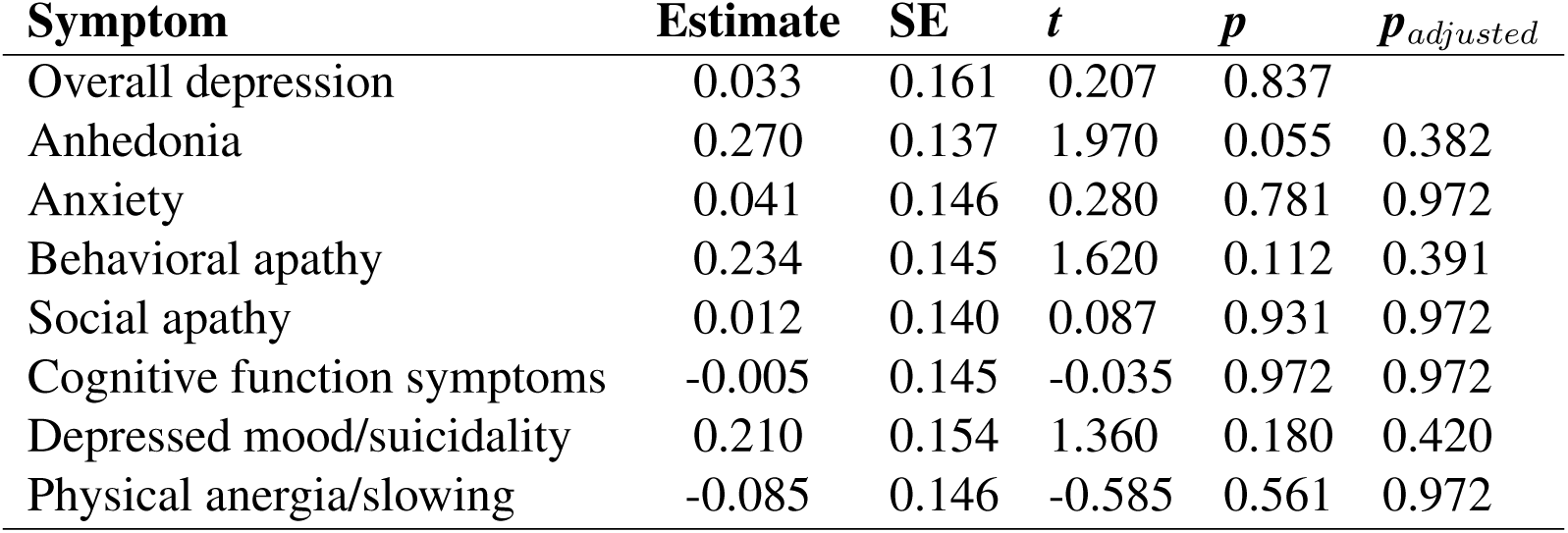
Symptoms inverse temperature regressions, MDD only. Predict inverse temperature by overall depression severity and each symptom domain, controlling for age, and years of education (* indicates *p<*0.05, FDR correction within symptom models). All variables were scaled as input to the regressions.

**Table S.I. 11:** Travel task performance diagnostic group differences. Column 1: predicted variable in regression, column 2: predictor variable, column 3: regression estimate, column 4: standard error (SE), column 5: degrees of freedom, column 6: t-statistic, column 7: p-value.

| Predicted variable | Predictor variable | Estimate | SE | df | *t* | *p* |
| --- | --- | --- | --- | --- | --- | --- |
| N-Back missed trials (%) | Group | -0.021 | 0.495 | 77 | -0.042 | 0.966 |
| N-Back RT (log) | Group | -0.163 | 0.123 | 78.63 | -1.35 | 0.181 |
| N-Back RT (log) | Group*correct*N-Back | -0.197 | 0.084 | 4955 | -2.335 | 0.020 |
| N-Back RT (log) | Group*correct* target | -0.273 | 0.009 | 2450 | -3.206 | 0.001 |
| Accuracy (D') | Group | -0.008 | 0.222 | 113.6 | -0.034 | 0.973 |
| Accuracy (D') | Group*N-Back level | 0.117 | 0.202 | 77 | 0.577 | 0.566 |
| Required keypresses | Group | -2.00 | 3.640 | 77 | -0.550 | 0.584 |
| Completed presses (%) | Group | 0.272 | 0.970 | 101 | 0.281 | 0.780 |
| Completed presses (%) | Group*effort level | -2.379 | 0.675 | 3237.4 | -3.524 | 0.0004 |
| Completed larger presses | Group | -1.490 | 1.510 | 79.33 | -0.987 | 0.327 |
| Completed smaller presses | Group | -0.010 | 0.358 | 1650 | -0.027 | 0.978 |

**Table S.I. 12:** Overall exit threshold relationship to symptoms (current MDD group, and all participants). Predicting individual differences in overall exit thresholds (log, from low effort conditions) by overall depression severity, and each symptom domain, controlling for age and years of education (* indicates *p<*0.05, FDR correction within symptom models). All variables were scaled as input to the regressions.

| Symptom | Estimate | SE | *t* | *p* | *p* *adjusted* |
| --- | --- | --- | --- | --- | --- |
| **A. Overall exit threshold, current MDD** |  |  |  |  |  |
| Overall depression | -0.42 | 0.20 | -2.12 | 0.040* |  |
| Anhedonia | -0.24 | 0.16 | -1.49 | 0.145 | 0.203 |
| Anxiety | -0.31 | 0.16 | -1.93 | 0.061 | 0.147 |
| Behavioral apathy | -0.28 | 0.16 | -1.77 | 0.084 | 0.147 |
| Social apathy | -0.20 | 0.16 | -1.25 | 0.219 | 0.255 |
| Cognitive function symptoms | -0.16 | 0.17 | -0.93 | 0.356 | 0.356 |
| Depressed mood/suicidality | -0.44 | 0.16 | -2.66 | 0.011* | 0.079 |
| Physical anergia/slowness | -0.33 | 0.17 | -1.90 | 0.065 | 0.147 |
| **B. Overall exit threshold, all participants** |  |  |  |  |  |
| Anhedonia | -0.03 | 0.12 | -0.29 | 0.775 | 0.775 |
| Anxiety | -0.13 | 0.12 | -1.07 | 0.289 | 0.506 |
| Behavioral apathy | -0.14 | 0.12 | -1.17 | 0.247 | 0.506 |
| Social apathy | -0.05 | 0.12 | -0.41 | 0.687 | 0.775 |
| Cognitive function symptoms | -0.10 | 0.12 | -0.86 | 0.395 | 0.553 |
| Depressed mood/suicidality | -0.22 | 0.13 | -1.65 | 0.104 | 0.506 |
| Physical anergia/slowness | -0.13 | 0.12 | -1.08 | 0.285 | 0.506 |

## Notes

### Competing Interest Statement

The authors have declared no competing interest.

### Funding Statement

This work was supported by the New Jersey Alliance for Clinical and Translational Science (grant number UL1TR003017 awarded to LB, AK, JC), the New Jersey Health Foundation (grant number NJHF#PC158-23 awarded to LB, AK, JC), and the National Institutes of Health (LB, grant numbers T32MH065214, T32DA007261).

### Author Declarations

Institutional Review Board of Rutgers University gave ethical approval for this work

### Summary of Updates

Updated in response to reviewer comments. In preparing the revision we discovered an issue involving the coding of missed trials (which affected the model's expected reward on less than 2% of trials). We also took the opportunity to simplify the computation of expected reward in the MVT model, and recomputed all the results. This affected results numerically but did not affect the substantive pattern of results or conclusions.

